# Development and validation of the Symptom Burden Questionnaire™ for Long Covid: A Rasch analysis

**DOI:** 10.1101/2022.01.16.22269146

**Authors:** Sarah E Hughes, Shamil Haroon, Anuradhaa Subramanian, Christel McMullan, Olalekan L Aiyegbusi, Grace M Turner, Louise Jackson, Elin Haf Davies, Chris Frost, Gary McNamara, Gary Price, Karen Matthews, Jennifer Camaradou, Jane Ormerod, Anita Walker, Melanie J Calvert

## Abstract

**Objective:** To describe the development and initial validation of a novel patient-reported outcome measure of Long COVID symptom burden, the Symptom-Burden Questionnaire for Long COVID (SBQ™-LC).

**Method and Findings:** This multi-phase, prospective mixed-methods study took place between April and August 2021 in the United Kingdom (UK). A conceptual framework and initial item pool were developed from published systematic reviews. Further concept elicitation and content validation was undertaken with adults with lived experience (n = 13) and clinicians (n = 10), and face validity was confirmed by the Therapies for Long COVID Study Patient and Public Involvement group (n = 25). The draft SBQ™-LC was field tested by adults with self-reported Long COVID recruited via social media and international Long COVID support groups (n = 274). Thematic analysis of interview and survey transcripts established content validity and informed construction of the draft questionnaire. Rasch analysis of field test data guided item and scale refinement and provided evidence of the final SBQ™-LC’s measurement properties. The Rasch-derived SBQ™-LC is composed of 17 independent scales with promising psychometric properties. Respondents rate symptom burden during the past 7-days using a dichotomous response or 4-point rating scale. Each scale provides coverage of a different symptom domain and returns a summed raw score that may be converted to a linear (0 – 100) score. Higher scores represent higher symptom burden.

**Conclusions:** The SBQ™-LC is a comprehensive patient-reported assessment of Long COVID symptom burden developed using modern psychometric methods. It measures symptoms of Long COVID important to individuals with lived experience and may be used to evaluate the impact of interventions and inform best practice in clinical management.

## INTRODUCTION

Since its emergence in 2019, the global COVID-19 pandemic has caused more than 250 million infections and over five million deaths [1]. For many individuals, the infection is mild and short-lived; however, a proportion of those infected continue to experience, or go on to develop, long-term symptoms that persist beyond the acute phase of infection. These persistent symptoms are known collectively as “post-acute sequelae of COVID-19 (PACS)”, “post-acute COVID-19”, “post-COVID-19 syndrome”, or “Long COVID” [2].

Symptom burden may be defined as the “subjective, quantifiable prevalence, frequency, and severity of symptoms placing a physiologic burden on patients and producing multiple negative, physical, and emotional patient responses” [3]. The symptoms reported by individuals with Long COVID are heterogenous, affecting multiple organ systems with fatigue, dyspnea, and impaired concentration being among the most prevalent symptoms [4–6]. Symptoms may be persistent, cyclical, or episodic in their presentation and can pose a significant burden for affected individuals with negative consequences for work capability, functioning, and quality of life [7–9]. There is a growing body of research on the prevalence, incidence, co-occurrence, and persistence of Long COVID signs and symptoms [4,5,7,10–13]. To date, these data have largely been collected using bespoke, cross-sectional survey tools due, in part, to the limited availability of condition-specific, validated self-report instruments.

Patient-reported outcomes (PROs) are measures of health reported directly by the patient without amendment or interpretation by a clinician or anyone else [14]. Validated PRO instruments developed specifically for Long COVID that address the complex, multi-factorial nature of the condition are needed urgently to further understanding of Long COVID symptoms and underlying pathophysiology, support best practice in the clinical management of patients, and to evaluate the safety, effectiveness, acceptability, and tolerability of interventions [15–17]. Validated PRO instruments have recently been developed to measure the global impact of Long COVID and there are several unvalidated screening tools, surveys, and questionnaires also available [18,19]. However, individuals living with Long COVID have suggested that existing self-report measures fail to capture the breadth of experienced symptoms [9,20,21]. To address the need for a comprehensive measure of self-reported symptom burden specific to Long COVID, this study used Rasch analysis to develop and validate, in accordance with US Food and Drug Administration (FDA) guidance, a novel PRO instrument, the Symptom Burden Questionnaire™ for Long COVID (SBQ™-LC) [14,22].

## METHODS

### Setting and study design

This multi-phase, prospective mixed-methods study (Fig 1) was nested within the Therapies for Long COVID for non-hospitalized individuals (TLC) study (ISRTCN Reference: 15674970) and took place from 14 April to 1 August 2021 [23]. A favorable ethical opinion was granted by the University of Birmingham Research Ethics Committee (Reference: ERN_21-0191).

**Figure 1.**
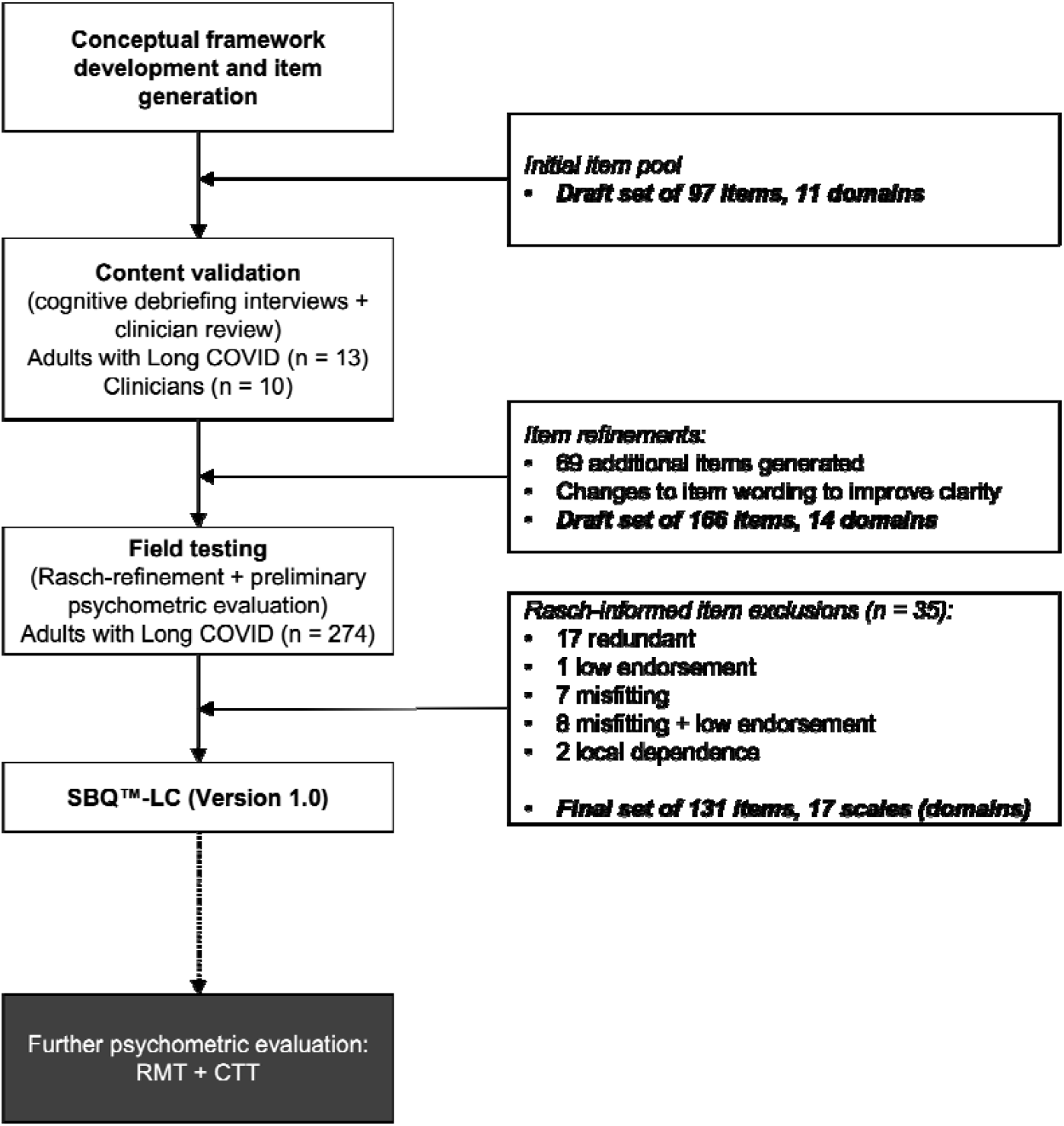
Study flow diagram showing the development of SBQ™-LC (Version 1.0)

### Study population

Content validation was undertaken with adults with Long COVID recruited from the TLC study’s Patient and Public Involvement and Engagement (PPIE) group and clinicians recruited from the TLC study team and Long COVID research studies based in the United Kingdom. The field test population included adults with self-reported Long COVID. Participants were aged 18 years or older who could self-complete the SBQ™-LC in English. No exclusion criteria relating to duration of Long COVID symptoms, hospitalization for SARS CoV-2, or vaccination status were applied. A minimum sample size of 250 respondents was prespecified for field testing. In Rasch analysis, a sample of 250 respondents provides 99% confidence that item calibrations and person measures are stable within ± 0.50 logits [24].

### Symptom coverage and existing PROs

The SBQ™-LC’s conceptual framework was developed from systematic literature reviews of Long COVID symptoms [4,25]. Existing symptom measures (n = 6) with good face validity in the context of Long COVID were reviewed to establish whether a new PRO instrument of symptom burden was needed [26–31]. When mapped to the conceptual framework, symptom coverage of these measures ranged from 25.0% to 64.1% (mean = 42.4% SD 16.8). This finding suggested complete coverage of Long COVID symptoms could not be guaranteed using existing measures, providing justification for the development of the SBQ™-LC.

### Study Procedures

#### Content validation

Content validation involved cognitive debriefing with adults with Long COVID and an online clinician survey. Cognitive debriefing interviews to ascertain the relevance, comprehensiveness, comprehensibility, and acceptability of the SBQ™-LC’s items for adults with lived experience of Long COVID were conducted via videoconferencing and recorded. The clinician survey was administered using the survey software application, SmartSurvey [32]. Verbatim transcripts of the interview recordings, field notes, and free text comments from the clinician survey were analyzed qualitatively using thematic analysis and a pre-specified framework to identify problems with the relevance, comprehensiveness, clarity, and acceptability of the items [33,34]. Participants with lived experience of Long COVID were asked to identify additional symptoms not captured by the initial item pool. Findings informed revisions which were tracked per item using an Excel spreadsheet. A draft version of the SBQ™-LC was constructed and sent forward for field testing.

#### Field testing

The Aparito Atom5™ platform is a regulated (ISO13485, ISO/IEC 27001:2013 Accreditation, FDA CFR21 Part 11 compliant) software platform that provides remote data collection and real-time patient monitoring via a smartphone application and integration with wearable devices [35]. The draft SBQ™-LC and a demographic questionnaire were programmed onto Atom5™ for delivery as an electronic patient-reported outcome (ePRO). Participants were recruited via social media advertisements posted on Twitter and Facebook by the study team and through other support group platforms and website registrations via collaborating Long COVID support groups based in the UK. Interested individuals clicked on a URL connecting them to a study-specific website where they could read detailed information about the study, provide their informed consent, and download the Atom5™ app to their mobile device (i.e., smartphone or tablet). Participants accessed the SBQ™-LC and demographic questionnaire via a unique QR code and once the completed questionnaires were submitted, participants could delete the app from their phone. The anonymised field test data were downloaded securely from Atom5™ for analysis.

### Statistical Analyses

STATA (Version 16) was used to clean and prepare the data and SPSS (Version 28.0) was used for descriptive data analyses. Rasch analysis was conducted on the field test data to refine the SBQ™-LC and assess its psychometric properties. A Rasch analysis is the formal evaluation of a PRO instrument against the Rasch measurement model. The Rasch model is a mathematical ideal that specifies a set of criteria for the construction of interval-level measures from ordinal data [36]. It is a probabilistic model that specifies that a person’s response to an item is only governed by the person and the location of the item on a shared scale measuring the latent trait. The probability that a person will endorse an item is a logistic function of the difference between a person’s trait level (expressed as person ability) and the amount of trait expressed by the item (expressed as item difficulty) [37]. Rasch analysis enabled the SBQ™-LC to be constructed as a modular PRO instrument (i.e., a multi-domain item bank) with linear, interval-level measurement properties. These properties render Rasch-developed PROs suitable for use with individual patients as well as for group-level comparisons, permit direct comparisons of scores across domains, and facilitate the construction of alternative test formats (i.e., short forms and computer adaptive tests (CATs) [38,39].

Rasch analyses were carried out using Winsteps software (Version 5.0.5) and the Partial Credit Model (PCM) for polytomous data. The PCM was selected as the question wording and rating scale categories varied across items. Joint Maximum Likelihood Estimation (JMLE) in Winsteps enabled parameter estimation where data were missing [40]. Items scored as “not applicable” were considered ‘missing’ and did not contribute to item estimates. Misfitting response patterns (e.g., arising from respondent guessing or other unexpected behaviour) have been shown to result in biased item estimates with detrimental impacts for model fit [41]. Therefore, as is customary in Rasch analysis, person fit statistics were appraised and persons with misfitting response patterns (i.e., outfit MnSq logits > 2.0 logits) were removed iteratively and item parameters re-estimated until evidence of item parameter stability was observed [37,42].

Rating scale functioning for individual items was assessed against the following criteria: 1) items oriented in the same direction as a check for data entry errors (i.e., appraisal of point-measure correlations), 2) average category measures advance (i.e., higher categories reflect higher measures; 3) category outfit mean-square (MnSq) values do not exceed 2.0 logits (i.e., as an indicator of unexpected randomness in the model); and 4) each category endorsed by a minimum of ten respondents [43]. If an item’s rating scale failed to meet these criteria, adjacent categories were combined or the item was removed. Category probability curves (CPCs) provided a graphical representation as further evidence of rating scale functioning.

To confirm model fit, Rasch analyses (including appraisal of unidimensionality, local independence, and individual item fit statistics) were completed iteratively as items were removed or grouped to create new scales. Person reliability and separation indices and scale-to-sample targeting were also evaluated. Targeting examines the correspondence between items and persons and, for a well-targeted scale, the items in a scale should be spaced evenly across a reasonable range of the scale and correspond to the range of the construct experienced by the sample. Person reliability examines the reproducibility of relative measure location and person separation provides a measure of the number of distinct levels of person ability (i.e., symptom burden) that can be distinguished by a scale [44]. Cronbach’s alpha, as a measure of the internal consistency reliability, was computed for each scale [45]. The list of parameters evaluated in the development and validation of the SBQ™-LC, along with acceptability criteria, are described in Table 1.

**Table 1.** Rasch measurement properties and acceptability criteria

| Measurement property | Definition/Aim of evaluation | Acceptability criteria for fit |
| --- | --- | --- |
| **Valid measurement model** | To identify a set of items which effectively measure the target construct of symptom burden in Long COVID (i.e., fulfil the axioms of fundamental measurement permitting the construction of interval level scales). | • Unidimensionality: PCAR<sup>a</sup> highest eigenvalue of first residual contrast < 2.0; disattenuated correlations > 0.70<br>• Local item independence: Residual correlation < 0.40<br>• Fit statistics: Outfit/infit MnSq values within 0.5 – 1.5 logits<br>• Point measure correlations > 0.40<br>• Rating scales: scale oriented with latent variable, categories advance monotonically, category fit statistics: Outfit MnSq values < 2.0 logits, uniform category endorsement. |
| **Reliability**<br>Rasch reliability | Rasch-based reliability is the share of true variance of the total observed variance of the measure. | • Rasch-based reliability:<br>$r \geq 0.70$ acceptable<br>$r \geq 0.80$ good<br>$r \geq 0.90$ excellent |
|  | Person separation index (PSI) is the number of distinct levels of person ability (i.e., symptom burden) that can be identified by the measure. | PSI<sup>b</sup> = 1.5 – 2.0 acceptable level of separation<br>PSI = 2.0 – 3.0 good level of separation<br>PSI ≥ 3.0 excellent level of separation |
| Internal consistency reliability | The extent to which items comprising a scale measure the same construct (i.e., the degree of homogeneity or relatedness among the items of a scale). | • Cronbach's alpha range $\geq 0.70$ |
| **Targeting** | The extent to which the range of the variable measured by the scale matches the range of that variable in the study sample. | • Item-person map<br>• Acceptable targeting is demonstrated by close correspondence of the person mean with the item mean for a scale (i.e., $\pm 1.0$ logits from the mean of zero). |

<sup>a</sup>Principal component analysis of the residuals; <sup>b</sup>PSI = person separation index

## RESULTS

### Item development and content validation

An initial item pool (n = 97) was constructed, guided by the conceptual framework developed from the published literature. Clinician review of the item pool informed changes to improve item clarity. Content validity was confirmed by individuals with lived experience of Long COVID (n = 13) in two rounds of cognitive debriefing interviews. Participants were of white ethnicity, ten (76.9%) were female, and ranged in age from 20 – 60 years. Cognitive debriefing identified gaps in symptom coverage, resulting in the generation of 69 new items. Thematic analysis classified problems with draft items’ relevance, comprehensiveness, clarity, acceptability. S1 Table presents key themes, together with exemplar quotations, from the thematic analysis.

The draft SBQ™-LC included 166 items (155 symptoms + 11 interference items) and an *a priori* theoretical domain classification comprised of 14 domains, each constructed as an independent scale. Items utilized a 7-day recall period and burden was measured using a dichotomous “yes/no” response or a 5-point rating scale measuring severity, frequency, or interference. Higher scores represented greater symptom burden. Face validity of the draft SBQ™-LC was confirmed by TLC study’s PPI group.

### Readability

Readability, measured using Flesch-Kincaid Reading Grade Level Test and the SMOG Index score, was calculated using the web-based application Readable [46–48]. The American Medical Association (AMA) and the National Institutes of Health (NIH) recommend readability of patient materials should not exceed a sixth-grade reading level (a Flesh-Kincaid Reading Grade Level of score of 6.0) [49]. The SBQ™-LC’s Flesch-Kincaid Grade Level score was 5.33 and SMOG index score was 8.27. Text with a SMOG index score ranging between 7 and 9 would be understood by 93% of adults in the UK [50].

### Field testing

#### Participants

Over a two-week period, 906 questionnaires were delivered and 330 responses received. Fifty-six submissions were incomplete and excluded from the analyses. The final sample included 274 complete responses, a response rate of 30.2%. Respondents’ ages ranged from 21 to 70 years, with a mean (SD) age of 45.1 (10.1) years. The sample included 240 (87.6%) females and 253 (92.3%) respondents were white. Two hundred and eleven (77.0%) participants reported three or more comorbidities. One hundred and twenty-nine (47.1%) participants reported a diagnosis of SARS-CoV-2 that was confirmed by a positive PCR test and 22 (8.0%) respondents reported confirmation of SARS-CoV-2 by a positive lateral flow test. Eleven participants (4.0%) had been hospitalized due to SARS-CoV-2 and 70 (25.5%) had received one dose of vaccine while 187 (68.2%) had received two doses. In total, 153 (55.8%) respondents were in either full-time or part-time employment (Table 2).

**Table 2.** Demographic characteristics of the field test sample (n = 274)

|  | n (%) |
| --- | --- |
| **Age (n = 263 reported)** |  |
| Mean | 45.1 years |
| SD | 10.1 years |
| Range | 21 – 70 years |
| **Gender** |  |
| Female | 240 (87.6) |
| Male | 34 (12.4) |
| **Ethnicity** |  |
| Asian or Asian British | 7 (2.6) |
| Black, African, Caribbean, or Black British | 3 (1.1) |
| Mixed or Multiple ethnic groups | 11 (4.0) |
| Other ethnic group | 0 (0.0) |
| White | 253 (92.3) |
| **Occupational status** |  |
| Caregiver | 3 (1.1) |
| Employed full-time | 114 (41.6) |
| Employed but currently not working | 51 (18.6) |
| Employed part-time | 39 (14.2) |
| Furloughed | 7 (2.6) |
| In full-time education | 3 (1.1) |
| Retired | 6 (2.2) |
| Voluntary work | 3 (1.1) |
| Unemployed | 20 (7.3) |
| Other | 28 (10.2) |
| **Positive PCR Test for SARS-Co-V-2 infection** |  |
| Yes | 129 (47.1) |
| No | 137 (50.0) |
| Don't know | 8 (2.9) |
| **Positive lateral flow test for SARS-Co-V-2 infection** |  |
| Yes | 22 (8.0) |
| No | 227 (82.8) |
| Don't know | 25 (9.1) |
| **Hospitalized for SAR-Co-V-2 infection** |  |
| Yes | 11 (4.0) |
| No | 263 (96.0) |
| **Admitted to intensive care unit due to SARS-Co-V-2 infection** |  |
| Yes | 0 (0.0) |
| No | 274 (100.0) |
| **Attended hospital emergency department due to SARS-Co-V-2 infection** |  |
| Yes | 111 (40.5) |
| No | 163 (59.5) |
| **Vaccine status as of (XX-Jun-2021)** |  |
| Two doses | 187 (68.2) |
| One dose | 70 (25.5) |
| No dose | 17 (6.2) |
| **Received shielding letter from UK government<sup>a</sup>** |  |
| Yes | 12 (4.4) |
| No | 262 (95.6) |
| **Care home resident** |  |
| Yes | 3 (1.1) |
| No | 271 (98.9) |
| **EQ5D utility index score (n = 262)** | 0.490 (SD 0.260) |

<sup>a</sup>Shielding letter = letter to indicate clinical vulnerability requiring enhanced social distancing.

#### Assessment of rating scale function, item and scale refinement

Rating scale functioning was assessed by examining the relevant Winsteps output tables and item category probability curves (S1 Figure). Appraisal of category endorsement revealed the presence of floor effects and a positively skewed distribution. To address non-uniform category distribution, the 5-point rating scale was collapsed to either a dichotomous response or 4-point rating scale (where 0 = none, 1 = mild, 2 = moderate, 3 = severe). After rating scale adjustment, point-measure correlations were found to be positive (range = 0.23 - 0.92), categories were ordered, and category fit statistics were productive for measurement (Outfit MnSq < 2.0 logits) for all items. Category distribution remained positively skewed (mean skewness = 1.44, mean SE = 0.15, range = -2.30 - 11.64). Disordered thresholds were observed for 52 (39.7%) items. Threshold disordering is indicative of a category occupying a narrow interval on the rating scale and is only considered a cause for concern if category disordering is also observed [40]. In total, 35 items were removed (Fig 1 shows the process of item reduction and reasons for removal). The remaining 131 items were systematically grouped to construct scales that were clinically sensible and satisfied the Rasch model.

#### Calibration of the final SBQ™-LC

Following response scale optimization and scale refinement, Rasch analyses were undertaken to report the psychometric properties of the final SBQ™-LC Version 1.0 (Table 3). The SBQ™-LC Version 1.0 is composed of 17 independent scales (Fig 2). All scales met the Rasch model criteria for unidimensionality and item fit. The first residual contrast values from principal component analyses of the residuals (PCAR) ranged from 1.46 to 2.03 eigenvalue units. No serious misfit was identified. Item infit MnSq values ranged from 0.67 to 1.32 logits and outfit MnSq values ranged from 0.44 to 1.53 logits. Fourteen item pairs showed local item dependency with residual correlation values > 0.4 (range = 0.44 – 0.88). In practical terms, a degree of local dependency is always observed in empirical data; therefore, it is necessary to consider the implications of item removal [51]. Despite evidence of local item dependence for eight scales, these items were retained to ensure comprehensive symptom coverage.

**Table 3.** Summary of Rasch analyses for the final SBQ™-LC (Version 1.0) (n = 274)

| Scale<br>(Symptom domain) | No<br>items | Misfitting<br>items | Persons<br>with<br>misfitting<br>responses<br>removed<br>(%) | Mean<br>person<br>ability<br>(logits) | PCA residual<br>eigenvalue<br>(1<sup>st</sup> contrast)<sup>a</sup> | Dependent<br>item pairs | Item<br>separation | Item<br>reliability | Person<br>separation<sup>b</sup> | Person<br>reliability<sup>c</sup> | Internal<br>consistency<br>reliability<sup>d</sup> |
| --- | --- | --- | --- | --- | --- | --- | --- | --- | --- | --- | --- |
| Breathing | 7 | 0 | 31 (11.3) | -1.95<br>(SE 0.99) | 1.72 | 0 | 7.68 | 0.98 | 1.8 | 0.76 | 0.84 |
| Circulation | 5 | 0 | 17 (6.2) | -1.16<br>(SE 0.98) | 1.58 | 0 | 7.43 | 0.98 | 1.1 | 0.55 | 0.63 |
| Fatigue | 4 | 0 | 33 (12.0) | 3.98<br>(SE 1.45) | 1.46 | 1 | 1.34 | 0.64 | 1.78 | 0.76 | 0.91 |
| Memory, Thinking &<br>Communication | 10 | 0 | 12 (4.38) | -1.04<br>(SE 0.54) | 1.76 | 0 | 12.22 | 0.99 | 2.58 | 0.87 | 0.9 |
| Sleep | 4 | 0 | 31 (11.3) | -0.10<br>(SE 0.78) | 1.62 | 2 | 13.44 | 0.99 | 1.67 | 0.74 | 0.56 |
| Movement | 3 | 0 | 31 (11.3) | -6.27<br>(SE 2.06) | 1.57 | 3 | 11.23 | 0.99 | 2.08 | 0.81 | 0.86 |
| Muscles & Joints | 9 | 0 | 23 (8.39) | -0.85<br>(SE 0.54) | 1.68 | 0 | 7.38 | 0.98 | 1.68 | 0.73 | 0.84 |
| Skin & Hair | 8 | 0 | 26 (9.49) | -2.38<br>(SE 0.92) | 1.55 | 0 | 4.6 | 0.95 | 0.71 | 0.34 | 0.68 |
| Eyes | 10 | 0 | 12 (4.38) | -1.21<br>(SE 0.72) | 1.85 | 0 | 4.28 | 0.95 | 1.08 | 0.54 | 0.72 |
| Ear, Nose & Throat | 14 | 0 | 11 (4.01) | -1.25<br>(SE 0.49) | 1.96 | 1 | 3.39 | 0.92 | 1.22 | 0.6 | 0.8 |
| Stomach & Digestion | 8 | 1 | 23 (8.39) | -1.52<br>(SE 0.70) | 1.6 | 0 | 6.19 | 0.97 | 0.95 | 0.48 | 0.7 |
| Mental Health &<br>Wellbeing | 9 | 0 | 25 (9.12) | -0.70<br>(SE 0.52) | 1.66 | 0 | 8.71 | 0.99 | 1.78 | 0.76 | 0.82 |
| Female Reproductive<br>& Sexual Health | 7 | 0 | 27 (9.85) | -2.07<br>(SE 1.05) | 1.99 | 2 | 5.52 | 0.97 | 0.99 | 0.5 | 0.59 |
| Male Reproductive &<br>Sexual Health | 3 | 0 | 1 (3.03) | -1.48<br>(SE 1.98) | 2.03 | 2 | 2.27 | 0.84 | 0.79 | 0.38 | 0.6 |
| Pain | 4 | 0 | 25 (9.12) | -1.53<br>(SE 1.05) | 1.75 | 2 | 13.06 | 0.99 | 1.62 | 0.72 | 0.77 |
| Other symptoms | 18 | 0 | 22 (8.03) | -1.53<br>(SE 0.43) | 1.67 | 1 | 5.5 | 0.97 | 1.33 | 0.64 | 0.79 |
| Interference | 8 | 0 | 21 (7.66) | 2.35<br>(SE 0.89) | 1.95 | 0 | 11.21 | 0.99 | 2.13 | 0.82 | 0.89 |

<sup>a</sup> Unidimensionality: PCAR eigenvalue of first residual contrast < 2.0; Person separation: $\geq 1.50$ acceptable, $\geq 2.00$ good, $\geq 3.00$ excellent; <sup>c</sup> Rasch-based reliability: $r \geq 0.70$ acceptable, $r \geq 0.80$ good, $r \geq 0.90$ excellent; <sup>d</sup> Cronbach's $\alpha \geq 0.70$ acceptable

**Figure 2.**
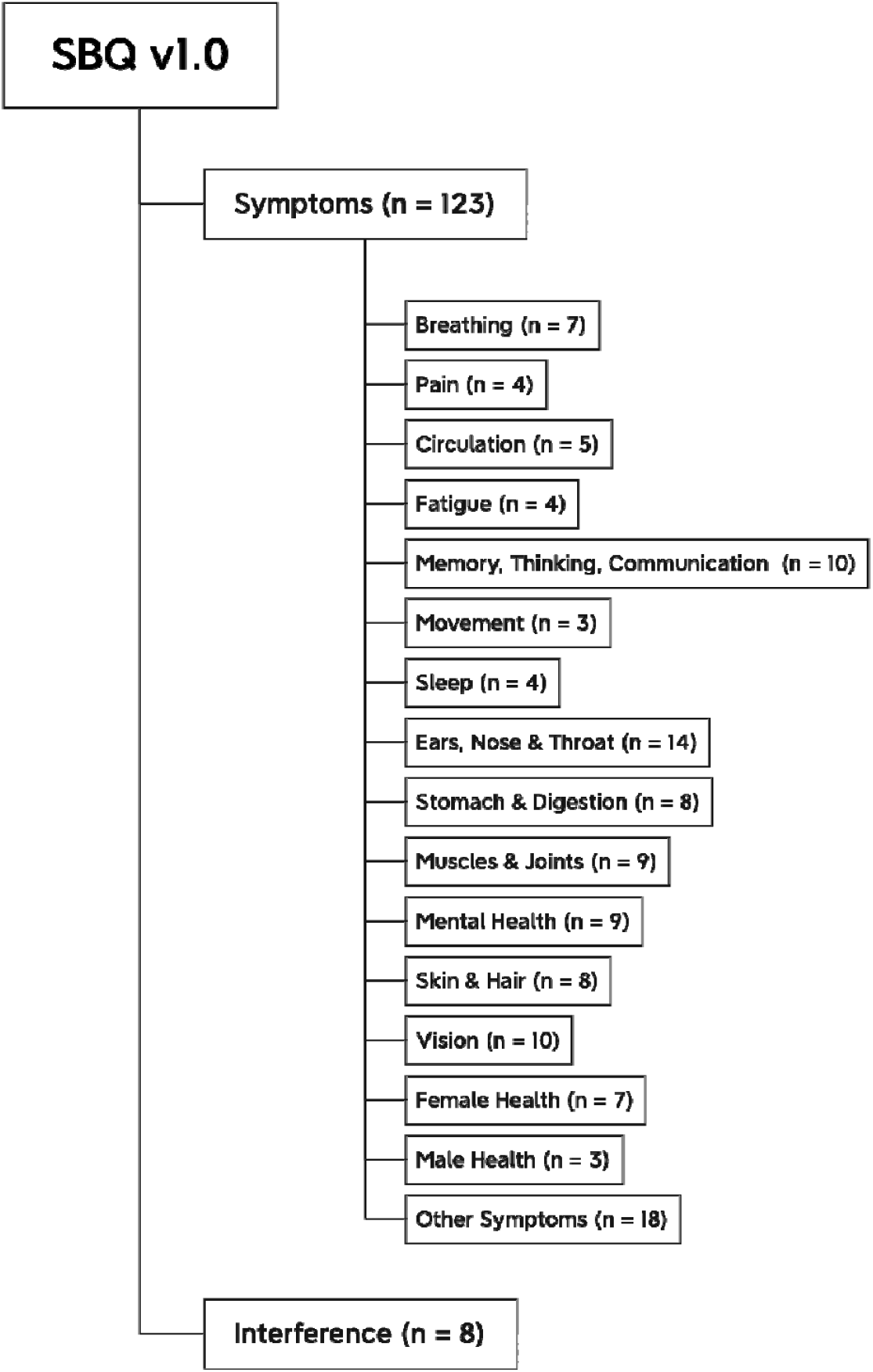
Conceptual framework of the final SBQ™-LC (Version 1.0)

Lastly, scale-to sample targeting and person reliability and separation were evaluated. Four scales had mean person ability values within ±1.0 logits of mean item difficulty. Mean person ability ranged from -6.27 logits to 3.98 logits (SE range = 0.43 - 2.06). S2 Figure shows the item-person maps for the SBQ™-LC’s scales. Person reliability ranged from 0.34 to 0.87 and person separation indices ranged from 0.71 – 2.58. Values for Cronbach’s alpha, as evidence of internal consistency reliability, ranged from 0.56 to 0.91.

## DISCUSSION

This study described the development and initial validation of the SBQ™-LC, a Rasch-developed, modular PRO instrument constructed to comprehensively assess symptom burden in Long COVID. The SBQ™-LC was developed in accordance with international, consensus-based standards and regulatory guidance and may be used to evaluate the impact of interventions and to inform clinical care [14,22,52]. The findings from published systematic reviews were used to construct a conceptual framework and generate an initial item pool. Rigorous content validity testing provided evidence of the SBQ™-LC’s relevance, comprehensiveness, comprehensibility, and acceptability. Rasch analysis permitted optimisation of the SBQ™-LC’s items and response scales.

The SBQ™-LC was developed with the extensive involvement of adults with lived experience of Long COVID and is a strength of this study. Involvement of the target population is regarded as the “gold standard” in the development of PRO instruments. Patient involvement may be considered of particular importance in the context of Long COVID where there is a rapidly evolving evidence-base and affected individuals have reported experiences of stigma and a lack of acknowledgement from the medical community regarding the breadth and nature of their symptoms [9]. Involvement of adults with Long COVID in all study phases (i.e., development, refinement, and validation) ensured the patient perspective was embodied in the SBQ™-LC’s items.

Rasch analysis of the SBQ™-LC guided item reduction and rating scale refinement, optimised measurement accuracy, and minimized respondent burden. These analyses showed several of the SBQ™-LC’s scales to be off-target with evidence of low person reliability and separation. Poor scale-to sample targeting is indicative of the items within a scale failing to provide full coverage of person locations (i.e., range of symptom burden experienced by the sample) [53]. Negative mean person measures (>1.0 logits), floor effects, and positively skewed response category distributions suggested the SBQ™-LC may be targeting individuals with higher levels of symptom burden than the level of burden represented by the field test sample. Highly skewed samples and poor targeting can produce low person reliability coefficients even if an instrument is functioning as intended and provides a possible explanation for the low person reliability values observed in this study [53].

As a Rasch-developed instrument, the SBQ™-LC’s ordinal raw scales may be converted to linear scales, with each 1-point change in a scale score being equidistant across the entire scale. Linear scores will enable the direct comparison of scores across the symptom domains represented by the SBQ™-LC’s scales. This modular construction means researchers and clinicians need only select those scales required to provide targeted assessment of a particular symptom domain, reducing respondent burden further by removing the need to complete the SBQ™-LC in its entirety. Moreover, the Rasch model will make it possible to compare data from the SBQ™-LC with other PROs via co-calibration studies. As each item of a Rasch-derived scale functions independently from others on that scale, the SBQ™-LC can be adapted for administration as a computer adaptive test (CAT) [39,54]. A CAT is a test, administered via a computer, which adapts to the respondent’s ability in real-time by selecting different questions from an item bank to provide a more accurate measure of the respondent’s ability without the need to administer a large number of items [55]. CATs can reduce respondent burden, improve accuracy, and provide individualized assessment, instrument characteristics which are attractive when assessing a health condition with heterogeneous, relapsing and remitting symptoms such as Long COVID.

The burden of Long COVID on healthcare systems continues to grow as more individuals are infected by SARS-CoV-2 [56]. To meet this growing demand, services require cost-effective resources to support safe, effective clinical management. The use of the SBQ™-LC in the TLC study will provide early evidence of feasibility of the SBQ™-LC for use in remote patient monitoring [57]. A previous randomized controlled trial has shown remote symptom monitoring using PROs can result in fewer emergency department attendances, reduce hospital admissions, prompt earlier intervention, and improve patients’ health-related quality of life [58]. If used in a clinical trial, symptom data collected remotely via the SBQ™-LC could provide valuable information on the safety, efficacy, and tolerability of new interventions for Long COVID [15]. If used within routine care, the SBQ™-LC has potential to facilitate patient-clinician conversations, guide treatment decision-making, and facilitate referrals to specialist services [58–60].

The use of social media to recruit participants meant it was not possible to confirm representativeness of the field test sample and is a limitation of this study. The relatively low completion rate suggested potential field test participants (i.e., possibly individuals experiencing higher levels of symptom burden, for example, symptoms such as fatigue or cognitive dysfunction) may have been deterred by the effort required to complete the SBQ™-LC or lacked sufficient incentive to participate. As such, the level of symptom burden captured in the field test may not reflect accurately the wider experience of Long COVID patients. Lastly, despite attempts to sample purposively for maximum variation, the demographics of the content validation study sample were highly skewed towards white (100%) females (76.9%). Further validation, undertaken as part of the TLC study which will prospectively target higher representation from ethnic minority groups, is planned and aims to confirm these findings. Further research is also needed to evaluate the SBQ™-LC using traditional psychometric indicators including test-retest reliability, construct validity, responsiveness, and measurement error. Studies to explore the feasibility and acceptability of the SBQ™-LC for use in clinical settings are also needed. The SBQ™-LC is currently available in UK English as an ePRO and in paper form. Linguistic and cross-cultural validation studies will ensure its generalizability outside of the UK and make translated versions of the SBQ™-LC available for use.

The presence of symptoms of COVID-19 persisting beyond the acute phase of infection in a significant number of patients represents an ongoing challenge for healthcare systems globally. High quality PRO instruments with robust psychometric properties are required to better understand the signs, symptoms, and underlying pathophysiology of Long COVID, to develop safe and effective interventions, and to meet the day-to-day needs of this growing patient group. The SBQ™-LC was developed as a comprehensive measure of Long COVID symptom burden. With promising psychometric properties, the SBQ™-LC may be suitable for use in Long COVID research studies and in the delivery of clinical care.

## Data Availability

Data for this project are not currently available for access outside the TLC research team. The dataset may be shared when finalised but this will require an application
to the data controllers. The data may then be released to a specific research team for a specific project dependent on the independent approvals being in place.

## Acknowledgements

The authors gratefully acknowledge the contributions of the clinicians who participated in the online survey. We thank Dr Anita Slade for their support with interpretation of the Rasch analyses.

The SBQ™-LC Version 1.0 is available under license. To request a copy, please.

## Contributors

*Concept and design:* Calvert, Haroon, Hughes, Aiyegbusi, Frost, Davies

*Acquisition, analysis, or interpretation of data:* Hughes, Haroon, Calvert, Aiyegbusi, Matthews, Camaradou, Price, Ormerod, McMullan, Davies, Frost, McNamara

*Drafting of the manuscript:* Hughes, Calvert, Haroon, Submaranian

*Critical revision of the manuscript:* Hughes, Calvert, Haroon, Submaranian, McMullan, Aiyegbusi, Davies, Frost, McNamara, Matthews, Camaradou, Jackson, Turner, Price, Ormerod, Walker

*Statistical analysis:* Hughes, Submaranian, Jackson

*Obtained funding:* Haroon, Calvert, Hughes, Aiyegbusi, McMullan, Davies

*Administration, technical, or material support:* Frost, Davies, Submaranian, Jackson, McNamara Walker

## Data availability statement

Data for this project are not currently available for access outside the TLC research team. The dataset may be shared when finalised but this will require an application to the data controllers. The data may then be released to a specific research team for a specific project dependent on the independent approvals being in place.

## SUPPORTING INFORMATION CAPTIONS

**S1 Table. Themes and exemplar quotations from content validation of the SBQ™-LC item pool (cognitive debriefing and online survey)**

**S1 Figure. Item category probability curves for the final SBQ™-LC (Version 1.0)**

Each graph, the y-axis represents the expected probability of endorsement of any given category when a person responds to the item. The x-axis represents the person ability relative to the item difficulty, with origins set to 0. The scale is measured in logits. Locations to the right of ‘0’ represent greater and greater levels of symptom burden. Moving to the left of ‘0’ locations represent lower levels of symptom burden. Categories should advance in ascending order from left to right along the x-axis and each have a distinct peak (indicating that the category is the most probable (modal) category at that point on the latent variable). The crossover points between two curves are the equal probability points of thresholds.

**S2 Figure. Item-person maps for the 17 scales of the final SBQ™-LC (Version 1.0)**

For each scale, the person-item map displays the location of person abilities and item difficulties respectively along the same latent dimension (y-axis).

## REFERENCES

1. COVID-19 situation update worldwide, as of week 44, updated 11 November 2021. [cited 16 Nov 2021]. Available from: https://www.ecdc.europa.eu/en/geographical-distribution-2019-ncov-cases

2. National Institute for Health and Care Excellence (NICE). COVID-19 rapid guideline: managing the long-term effects of COVID-19. [cited 3 Dec 2021]. Available from: https://app.magicapp.org/#/guideline/EQpzKn/section/n3vwoL

3. Gapstur RL. Symptom Burden: A Concept Analysis and Implications for Oncology Nurses. Oncol Nurs Forum. 2007;34: 673–680. doi:10.1188/07.ONF.673-680. PMID: 17573326.

4. Aiyegbusi OL, Hughes SE, Turner G, Rivera SC, McMullan C, Chandan JS, Haroon S, Price G, Davies EH, Nirantharakumar K, Sapey E, Calvert MJ; TLC Study Group. Symptoms, complications and management of long COVID: a review. J R Soc Med. 2021 Sep;114(9):428–442. doi: 10.1177/01410768211032850. PMID: 34265229.

5. Cabrera Martimbianco AL, Pacheco RL, Bagattini ÂM, Riera R. Frequency, signs and symptoms, and criteria adopted for long COVID-19: A systematic review. Int J Clin Pract. 2021 Oct;75(10):e14357. doi: 10.1111/ijcp.14357. PMID: 33977626.

6. CDC. Post-COVID Conditions. In: Centers for Disease Control and Prevention [Internet]. 11 Feb 2020 [cited 7 Dec 2021]. Available from: https://www.cdc.gov/coronavirus/2019-ncov/long-term-effects/index.html

7. Amin-Chowdhury Z, Harris RJ, Aiano F, Zavala M, Bertran M, Borrow R, et al. Characterising long COVID more than 6 months after acute infection in adults; prospective longitudinal cohort study, England. medRxiv [Preprint]. 2021 Mar [cited 2022 Jan 14]. Available from: https://www.medrxiv.org/content/10.1101/2021.12.20.21268098v1 doi:10.1101/2021.03.18.21253633

8. Parker AM, Brigham E, Connolly B, McPeake J, Agranovich AV, Kenes MT, Casey K, Reynolds C, Schmidt KFR, Kim SY, Kaplin A, Sevin CM, Brodsky MB, Turnbull AE. Addressing the post-acute sequelae of SARS-CoV-2 infection: a multidisciplinary model of care. Lancet Respir Med. 2021 Nov;9(11):1328–1341. doi: 10.1016/S2213-2600(21)00385-4. PMID: 34678213.

9. Ladds E, Rushforth A, Wieringa S, Taylor S, Rayner C, Husain L, Greenhalgh T. Persistent symptoms after Covid-19: qualitative study of 114 “long Covid” patients and draft quality principles for services. BMC Health Serv Res. 2020 Dec 20;20(1):1144. doi: 10.1186/s12913-020-06001-y. PMID: 33342437.

10. Taquet M, Dercon Q, Luciano S, Geddes JR, Husain M, Harrison PJ. Incidence, co-occurrence, and evolution of long-COVID features: A 6-month retrospective cohort study of 273,618 survivors of COVID-19. PLoS Med. 2021 Sep 28;18(9):e1003773. doi: 10.1371/journal.pmed.1003773. PMID: 34582441.

11. Sudre CH, Murray B, Varsavsky T, Graham MS, Penfold RS, Bowyer RC, et al. Attributes and predictors of long COVID. Nat Med. 2021;27(4): 626–631. doi:10.1038/s41591-021-01292-y. PMID: 33692530.

12. Nasserie T, Hittle M, Goodman SN. Assessment of the Frequency and Variety of Persistent Symptoms Among Patients With COVID-19: A Systematic Review. JAMA Netw Open. 2021 May 3;4(5):e2111417. doi: 10.1001/jamanetworkopen.2021.11417. PMID: 34037731.

13. Sudre CH, Murray B, Varsavsky T, Graham MS, Penfold RS, Bowyer RC, et al. Author Correction: Attributes and predictors of long COVID. Nat Med. 2021 Jun;27(6):1116. doi: 10.1038/s41591-021-01361-2. Erratum for: Nat Med. 2021 Apr;27(4):626-631. PMID: 34045738

14. FDA. Guidance for Industry Use in Medical Product Development to Support Labeling Claims Guidance for Industry. Clinical/Medical Federal Register. 2009; 1–39. doi:10.1111/j.1524-4733.2009.00609.x

15. Aiyegbusi OL, Calvert MJ. Patient-reported outcomes: central to the management of COVID-19. Lancet. 2020 Aug 22;396(10250):531. doi: 10.1016/S0140-6736(20)31724-4. Epub 2020 Aug 10. PMID: 32791038.

16. Berry P. Use patient reported outcome measures (PROMs) in treatment of long covid. BMJ. 2021 May 18;373:n1260. doi: 10.1136/bmj.n1260. PMID: 34006526.

17. Calvert M, Brundage M, Jacobsen PB, Schünemann HJ, Efficace F. The CONSORT Patient-Reported Outcome (PRO) extension: implications for clinical trials and practice. Health Qual Life Outcomes. 2013 Oct 29;11:184. doi: 10.1186/1477-7525-11-184. PMID: 24168680.

18. Tran VT, Riveros C, Clepier B, Desvarieux M, Collet C, Yordanov Y, et al. Development and validation of the long covid symptom and impact tools, a set of patient-reported instruments constructed from patients’ lived experience. Clin Infect Dis. 2021 Apr 29:ciab352. doi: 10.1093/cid/ciab352. Epub ahead of print. PMID: 33912905.

19. O’Connor RJ, Preston N, Parkin A, Makower S, Ross D, Gee J, et al. The COVID-19 Yorkshire Rehabilitation Scale (C19-YRS): Application and psychometric analysis in a post-COVID-19 syndrome cohort. J Med Virol. 2021 Oct 21:10.1002/jmv.27415. doi: 10.1002/jmv.27415. PMID: 34676578.

20. Alwan NA. The teachings of Long COVID. Commun Med. 2021;1: 1–3. doi:10.1038/s43856-021-00016-0

21. Callard F, Perego E. How and why patients made Long Covid. Soc Sci Med. 2021 Jan;268:113426. doi: 10.1016/j.socscimed.2020.113426. PMID: 33199035.

22. FDA. Principles for Selecting, Developing, Modifying, and Adapting Patient-Reported Outcome Instruments for Use in Medical Device Evaluation: Draft Guidance for Industry and Food and Drug Administration Staff, And Other Stakeholders. 2020 Aug [cited 2022 Jan 14]. Available from: https://www.fda.gov/regulatory-information/search-fda-guidance-documents/principles-selecting-developing-modifying-and-adapting-patient-reported-outcome-instruments-use

23. Haroon S, Nirantharakumar K, Hughes SE, Subramanian A, Aiyegbusi OL, Davies EH, et al. Therapies for Long COVID in non-hospitalised individuals - from symptoms, patient-reported outcomes, and immunology to targeted therapies (The TLC Study): Study protocol. medRxiv [Preprint]. 2021 [cited 14 Jan 2022]. Available from: https://www.medrxiv.org/content/10.1101/2021.12.20.21268098v1 doi:10.1101/2021.12.20.21268098

24. Linacre JM. Sample Size and Item Calibration or Person Measure Stability. Rasch Measurement Transactions. 1994;7: 328.[Cited 2022 Jan 14]. Available from: https://www.rasch.org/rmt/rmt74m.htm

25. Lopez-Leon S, Wegman-Ostrosky T, Perelman C, Sepulveda R, Rebolledo PA, Cuapio A, et al. More than 50 long-term effects of COVID-19: a systematic review and meta-analysis. Sci Rep. 2021 Aug 9;11(1):16144. doi: 10.1038/s41598-021-95565-8. PMID: 34373540.

26. de Haes JC, van Knippenberg FC, Neijt JP. Measuring psychological and physical distress in cancer patients: structure and application of the Rotterdam Symptom Checklist. Br J Cancer. 1990 Dec;62(6):1034–8. doi: 10.1038/bjc.1990.434. PMID: 2257209.

27. Basch E, Reeve BB, Mitchell SA, Clauser SB, Minasian LM, Dueck AC, et al. Development of the National Cancer Institute’s patient-reported outcomes version of the common terminology criteria for adverse events (PRO-CTCAE). J Natl Cancer Inst. 2014 Sep 29;106(9):dju244. doi: 10.1093/jnci/dju244. PMID: 25265940.

28. Cleeland CS, Mendoza TR, Wang XS, Chou C, Harle MT, Morrissey M, et al. Assessing symptom distress in cancer patients: the M.D. Anderson Symptom Inventory. Cancer. 2000 Oct 1;89(7):1634–46. doi: 10.1002/1097-0142(20001001)89:7<1634::aid-cncr29>3.0.co;2-v. PMID: 11013380.

29. Powers JH, Guerrero ML, Leidy NK, Fairchok MP, Rosenberg A, Hernández A, et al. Development of the Flu-PRO: a patient-reported outcome (PRO) instrument to evaluate symptoms of influenza. BMC Infect Dis. 2016 Jan 5;16:1. doi: 10.1186/s12879-015-1330-0. PMID: 26729246.

30. Evans RA, McAuley H, Harrison EM, Shikotra A, Singapuri A, Sereno M, et al. Physical, cognitive, and mental health impacts of COVID-19 after hospitalisation (PHOSP-COVID): a UK multicentre, prospective cohort study. Lancet Respir Med. 2021;9: 1275–1287. doi:10.1016/S2213-2600(21)00383-0. PMID: 34627560.

31. Smart Survey. Available from: https://www.smartsurvey.co.uk/

32. Braun V, Clarke V. Using thematic analysis in psychology. Qual Res Psychol. 2006;3: 77–101.

33. Willis GB. Analysis of the Cognitive Interview in Questionnaire Design. Oxford: Oxford University Press; 2015.

34. Aparito Limited [Cited 2022 Jan 14]. Available from: https://www.aparito.com/

35. Rasch G. Probabilistic Models for Some Intelligence and Attainment Tests. Copenhagen: Nielson and Lydiche; 1960. doi:10.1016/S0019-9958(61)80061-2

36. Bond TG, Yan Z, Heene M. Applying the Rasch Model: Fundamental Measurement in the Human Sciences. 4th edition. Routledge; 2020.

37. Hobart J, Cano S. Improving the evaluation of therapeutic interventions in multiple sclerosis: The role of new psychometric methods. Health Technol Assess. 2009;13. doi:10.3310/hta13120. PMID: 19216837.

38. Tennant A, McKenna SP, Hagell P. Application of Rasch analysis in the development and application of quality-of-life instruments. Value Health. 2004;7: S22–S26. doi:10.1111/j.1524-4733.2004.7s106.x. PMID:15367240.

39. Linacre JM. A User’s Guide to WINSTEPS MINISTEPS Rasch-Model Computer Programs. 2020. doi:ISBN 0-941938-03-4

40. Mousavi A, Cui Y. The Effect of Person Misfit on Item Parameter Estimation and Classification Accuracy: A Simulation Study. Education Sciences. 2020;10: 324. doi:10.3390/educsci10110324

41. Tennant A, Conaghan PG. The Rasch measurement model in rheumatology: What is it and why use it? When should it be applied, and what should one look for in a Rasch paper? Arthritis Rheum. 2007. doi:10.1002/art.23108. PMID: 18050173.

42. Linacre J. Optimizing Rating Scale Category Effectiveness. J Appl Meas. 2002;3(1): 85–106. PMID:11997586.

43. Duncan PW, Bode RK, Lai SM, Perera S. Rasch analysis of a new stroke-specific outcome scale: The stroke impact scale. Arch Phys Med Rehabil. 2003;84: 950–963. doi:10.1016/S0003-9993(03)00035-2. PMID: 12881816.

44. Streiner DL, Norman GR, Cairney J. Health Measurement Scales: A practical guide to their development and use. Health Measurement Scales. Oxford University Press; 2015.

45. Flesch R. A new readability yardstick. J Applied Psychol. 1948;32(3): 221–233. doi:10.1037/h0057532. PMID: 18867058.

46. McLaughlin GH. SMOG grading: A new readability formula. Journal of Reading. 1969;12: 639–646.

47. Readable. Available: http://www.readable.io

48. Weiss BD. Help patients understand: Manual for clinicians. American Medical Association Foundation and American Medical Association; 2007 May [cited 2022 Jan 14]. Available from: https://psnet.ahrq.gov/issue/health-literacy-and-patient-safety-help-patients-understand-manual-clinicians-2nd-ed

49. NHS Digital Service Manual. Use a readability tool to prioritise content - NHS digital service manual. In: nhs.uk [Internet]. Jul 2019 [cited 22 Jul 2020]. Available from: https://service-manual.nhs.uk

50. Local Dependency and Rasch Measures. [cited 10 Dec 2021]. Available from: https://www.rasch.org/rmt/rmt213b.htm

51. Mokkink LB, de Vet HCW, Prinsen CAC, Patrick DL, Alonso J, Bouter LM, Terwee CB. COSMIN Risk of Bias checklist for systematic reviews of Patient-Reported Outcome Measures. Qual Life Res. 2018 May;27(5):1171–1179. doi: 10.1007/s11136-017-1765-4. PMID: 29260445;

52. Separation, Reliability and Skewed Distributions: Statistically Different Levels of Performance. [cited 6 Dec 2021]. Available from: https://www.rasch.org/rmt/rmt144k.htm

53. Harrison CJ, Geerards D, Ottenhof MJ, Klassen AF, Riff KWYW, Swan MC, et al. Computerised adaptive testing accurately predicts CLEFT-Q scores by selecting fewer, more patient-focused questions. J Plast Reconstr Aesthet Surg. 2019. doi:10.1016/j.bjps.2019.05.039. PMID: 31358447.

54. Computer Adaptive Testing: Background, benefits and case study of a large-scale national testing programme. In: Surpass, Powering Assessment [Internet]. 11 Dec 2019 [cited 5 Dec 2021]. Available from: https://surpass.com/news/2019/computer-adaptive-testing-background-benefits-and-case-study-of-a-large-scale-national-testing-programme/

55. Menges D, Ballouz T, Anagnostopoulos A, Aschmann HE, Domenghino A, Fehr JS, et al. Burden of post-COVID-19 syndrome and implications for healthcare service planning: A population-based cohort study. PLOS ONE. 2021;16: e0254523. doi:10.1371/journal.pone.0254523. PMID: 34252157

56. Vindrola-Padros C, Sidhu MS, Georghiou T, Sherlaw-Johnson C, Singh KE, Tomini SM, et al. The implementation of remote home monitoring models during the COVID-19 pandemic in England. EClinicalMedicine. 2021;34. doi:10.1016/j.eclinm.2021.100799. PMID: 33817610.

57. Basch E, Deal AM, Kris MG, Scher HI, Hudis CA, Sabbatini P, et al. Symptom monitoring with patient-reported outcomes during routine cancer treatment: A randomized controlled trial. J Clin Oncol. 2016;34: 557–565. doi:10.1200/JCO.2015.63.0830. PMID: 26644527.

58. Greenhalgh J, Gooding K, Gibbons E, Dalkin S, Wright J, Valderas J, et al. How do patient reported outcome measures (PROMs) support clinician-patient communication and patient care? A realist synthesis. J Patient Rep Outcomes. 2018;2. doi:10.1186/s41687-018-0061-6. PMID: 30294712.

59. Calvert M, Kyte D, Price G, Valderas JM, Henrik Hjollund N. Maximising the impact of patient reported outcome assessment for patients and society. BMJ. 2019;364. doi:10.1136/bmj.k5267. PMID: 30679170.

