## Supplementary material for "Development and validation of the Symptom Burden Questionnaire™ for Long Covid: A Rasch analysis": S1 Table

**S1 Table: Themes and exemplar quotations from content validation of the SBQ™-LC item pool (cognitive debriefing and online survey)**

| Theme | Exemplar quote |
|---|---|
| Content relevance | *"...The only the only question that I sort of went .... was number seven around how often did you feel like you were like the person you were before your illness. And I think that's probably the most important [question]...yeah, that was the most emotive one for me...."* – Participant 02 |
| Content comprehensiveness | *"...the questions are unique in the way they capture, show understanding of, and validate the range of symptoms a person with Long COVID can experience..."* – Participant 12<br><br>*I think you really need a section for neuro, neurological aspects in there. And the eyes, bloodshot eyes, and things like that but otherwise I think you, you kind of captured most of the symptoms, if I think.* -Participant 10<br><br>*"No, I think I think it is really good that it's structured this way and it's been detailed in terms of the shortness at the breath, fatigue, the muscle and joint pain. Because even me, I couldn't describe although I feel them, I don't have a medical knowledge. Expanding and going into details, I think it's really good, I think it is really helpful for whoever is going to take the questionnaire. It's very helpful."* – Participant 13 |
| Item clarity | *"...the difficulty is around the fluctuation and the seven-day window. For example, breathlessness is a big one for me. I can go weeks and feel ok, and then, all of a sudden, it relapses and I'm struggling to breathe again"* - Participant 02<br><br>*"...'Fatigue', to me, is where I feel like I need to sleep. I'm probably sleeping between 10 and 11 hours and the moment and then napping in the day. I think 'low energy' is kind of having that draining feeling."* – Participant 01 |
| Item acceptability | *"[I] thought it was relatively straightforward, I would say. My concentration isn't good, and I managed."* - Participant 12 |
| Response scales | *"...the difficulty is around the fluctuation and the seven-day window. For example, breathlessness is a big one for me. I can go weeks and feel ok, and then, all of a sudden, it relapses and I'm struggling to breathe again"* - Participant 02 |
