## Supplementary material for "Development and validation of the Symptom Burden Questionnaire™ for Long Covid: A Rasch analysis": S1 Figure

#### Category Probability Curves – Breathing

For each graph, the y-axis represents the expected probability of endorsement of any given category when a person responds to the item. The x-axis represents the person ability relative to the item difficulty, with origins set to 0. The scale is measured in logits. Locations to the right of ‘0’ represent greater and greater levels of symptom burden. Moving to the left of ‘0’ locations represent lower levels of symptom burden. Categories should advance in ascending order from left to right along the x-axis and each have a distinct peak (indicating that the category is the most probable (modal) category at that point on the latent variable). The crossover points between two curves are the equal probability points of thresholds. Each category curved is colour coded: category 0 = red, category 1 = blue; category 2 = pink; category 3-4 = dark green.

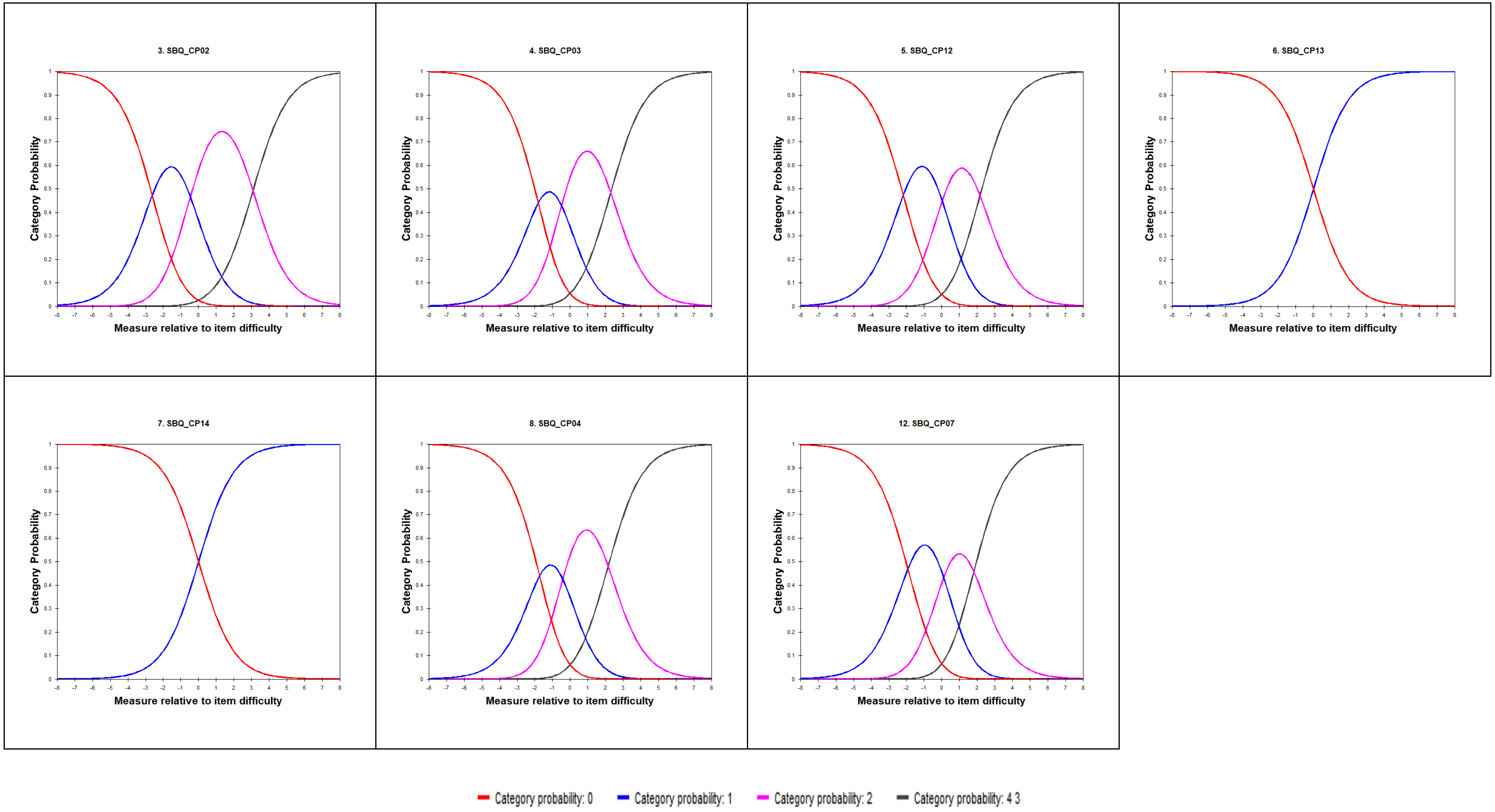
3. SBQ_CP02
4. SBQ_CP03
5. SBQ_CP12
6. SBQ_CP13
7. SBQ_CP14
8. SBQ_CP04
12. SBQ_CP07
Category Probability
Measure relative to item difficulty
Category probability: 0
Category probability: 1
Category probability: 2
Category probability: 4 3

### Category Probability Curves – Pain

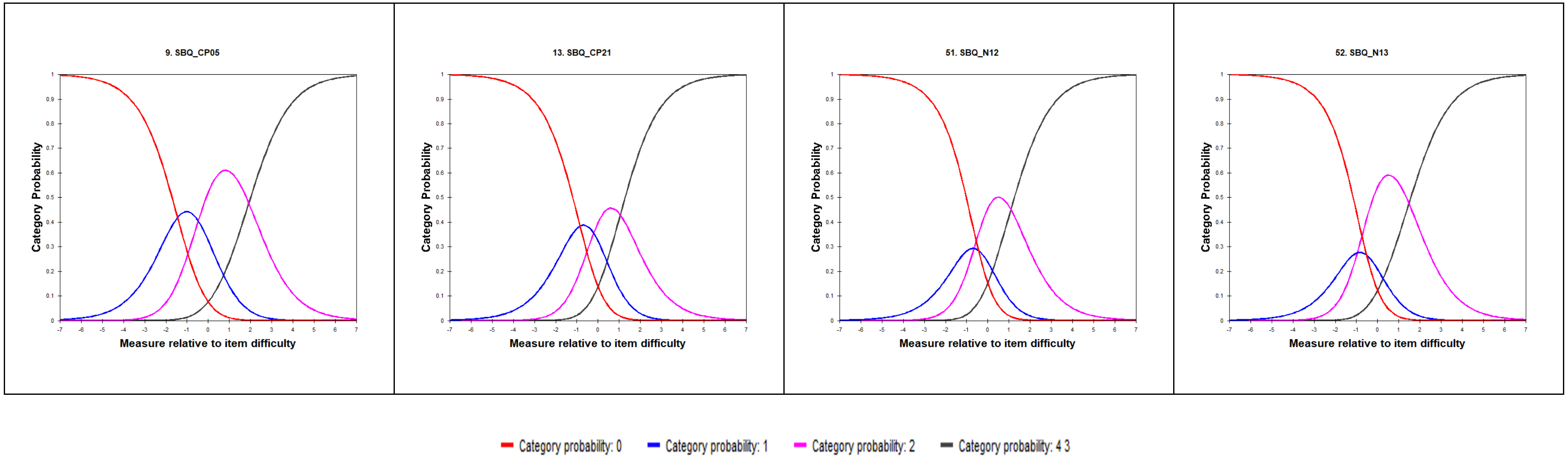

### Category Probability Curves – Circulation

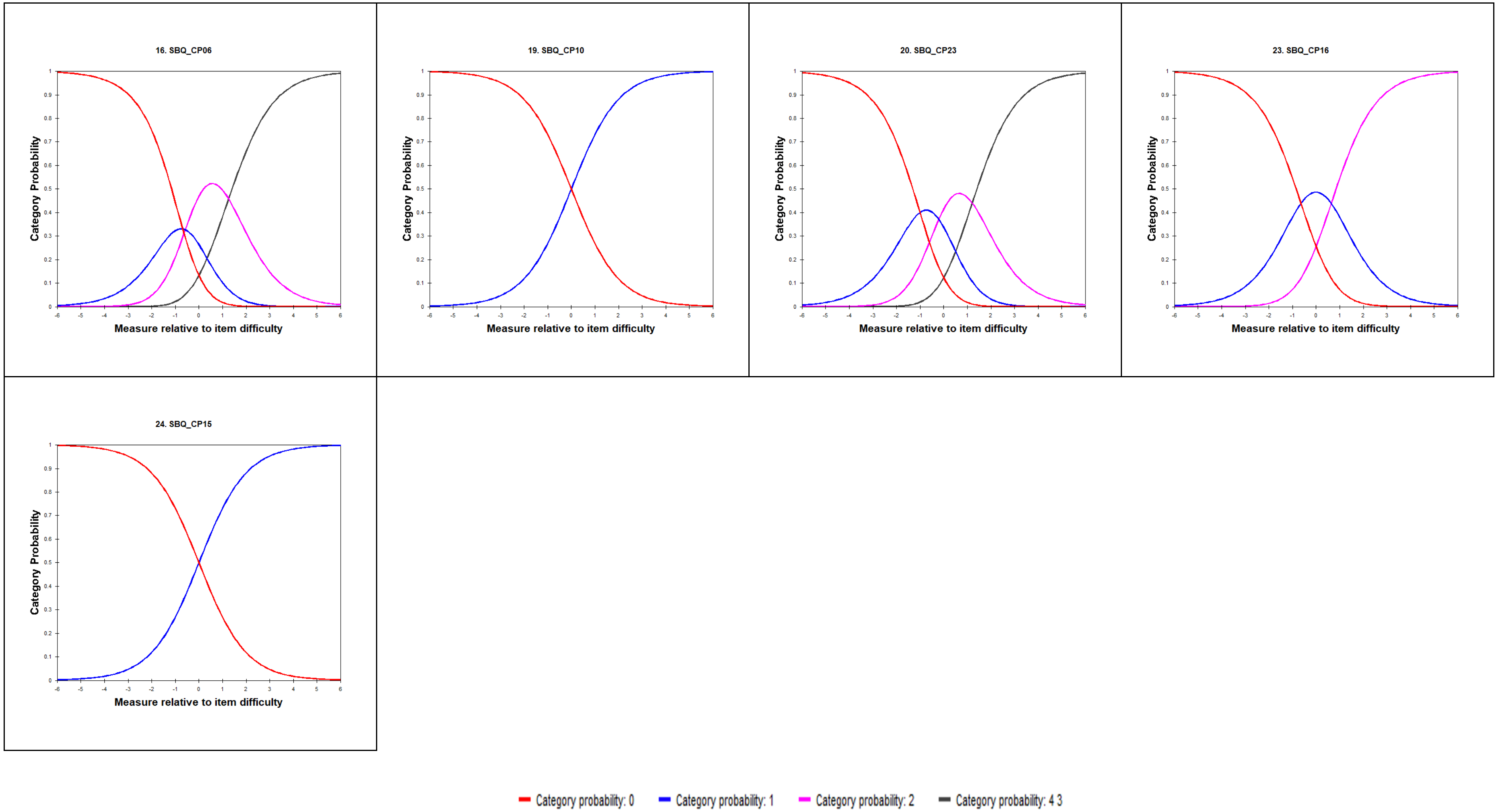

#### Category Probability Curves – Fatigue

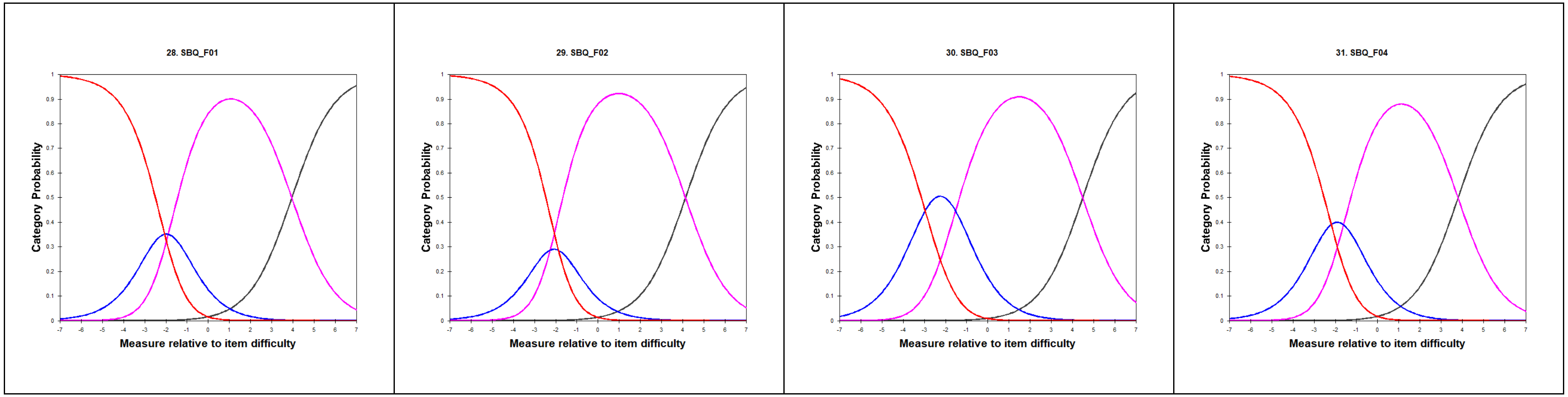


Category probability: 0 Category probability: 1 Category probability: 2 Category probability: 4 3

### Category Probability Curves – Memory, Thinking and Communication

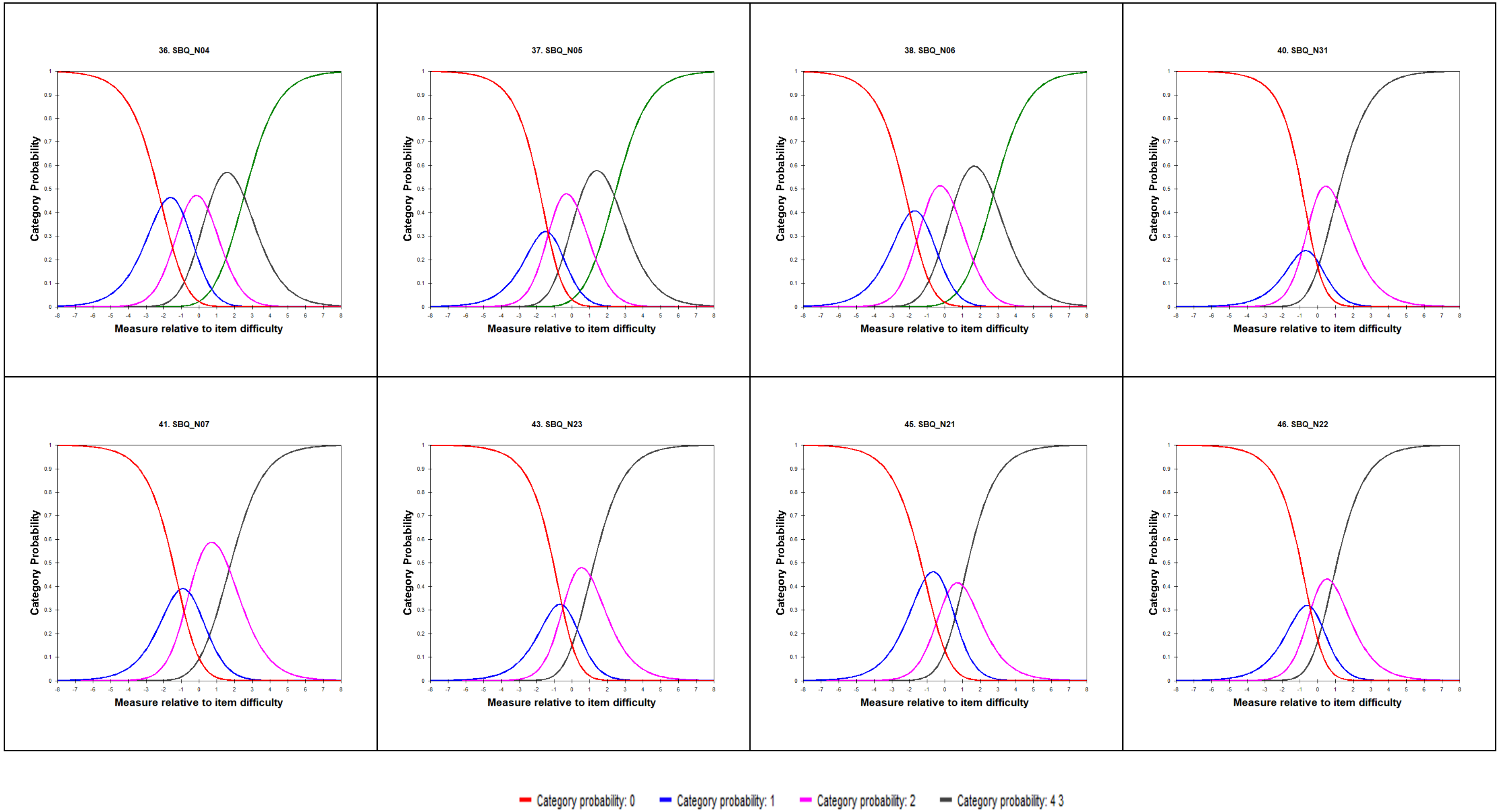

#### Category Probability Curves – Memory, Thinking and Communication continued...

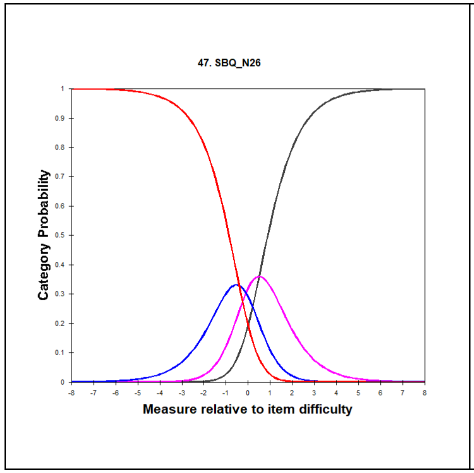


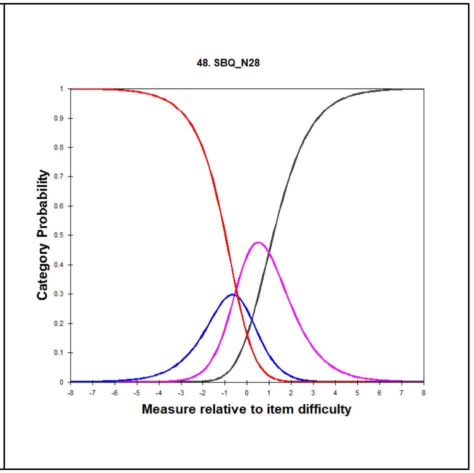


Category probability: 0 — Category probability: 1 — Category probability: 2 — Category probability: 4 3

#### Category Probability Curves – Movement

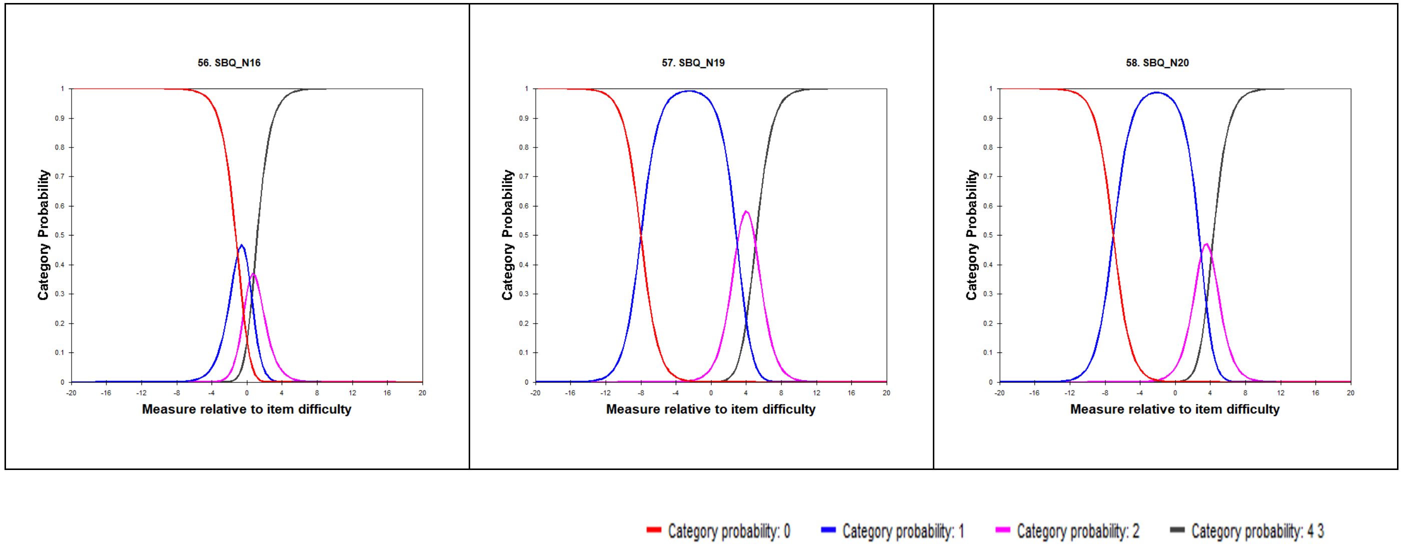

### Category Probability Curves – Sleep

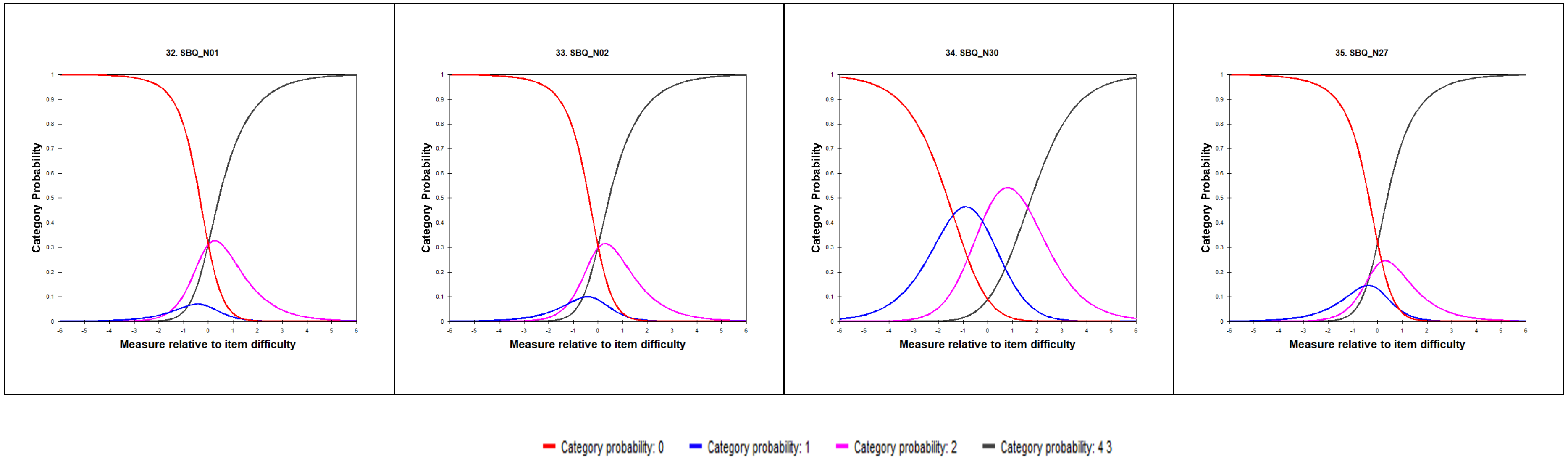

#### Category Probability Curves – Ear, Nose and Throat

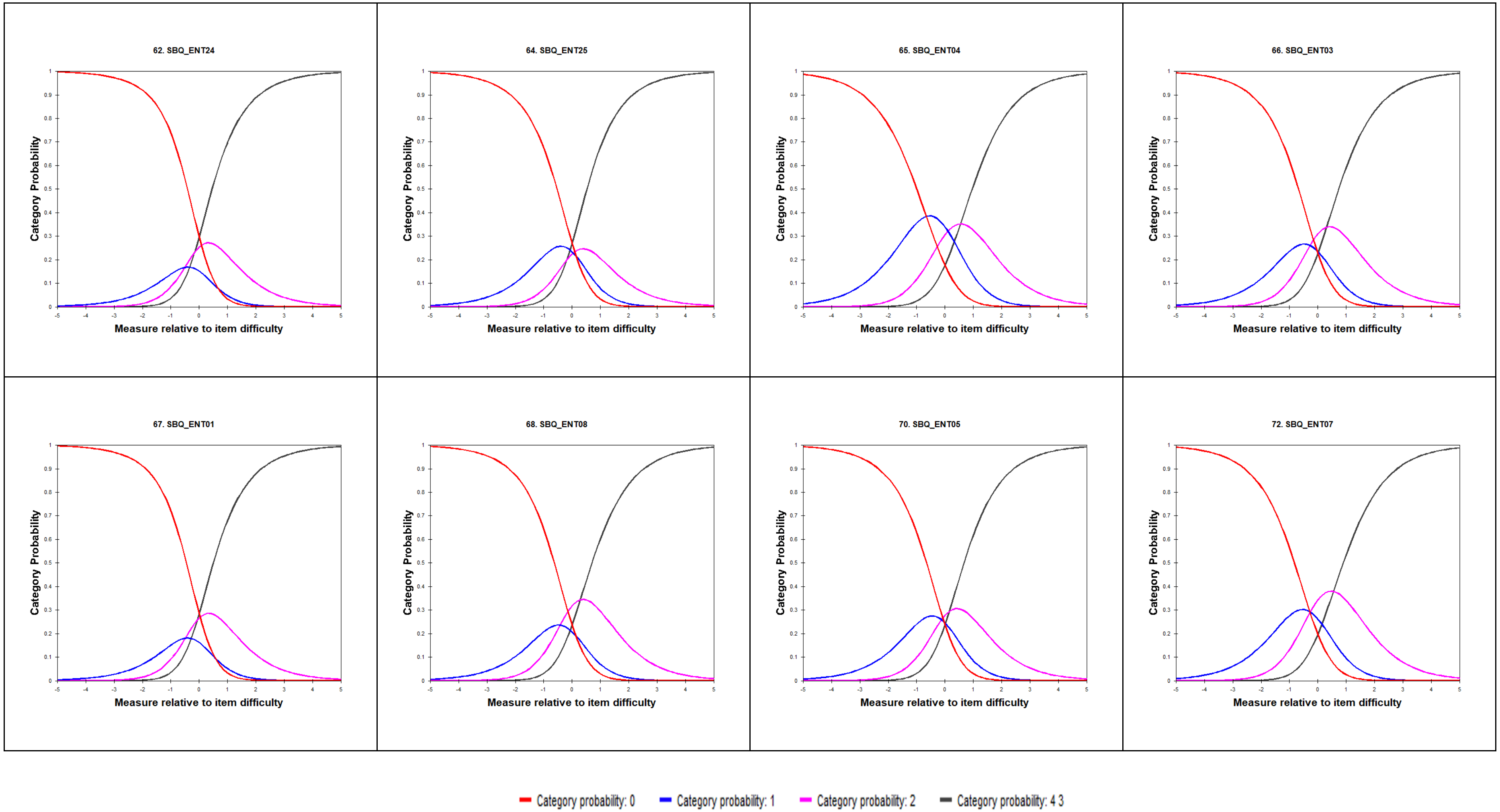

### Category Probability Curves: Ear, Nose and Throat continued....

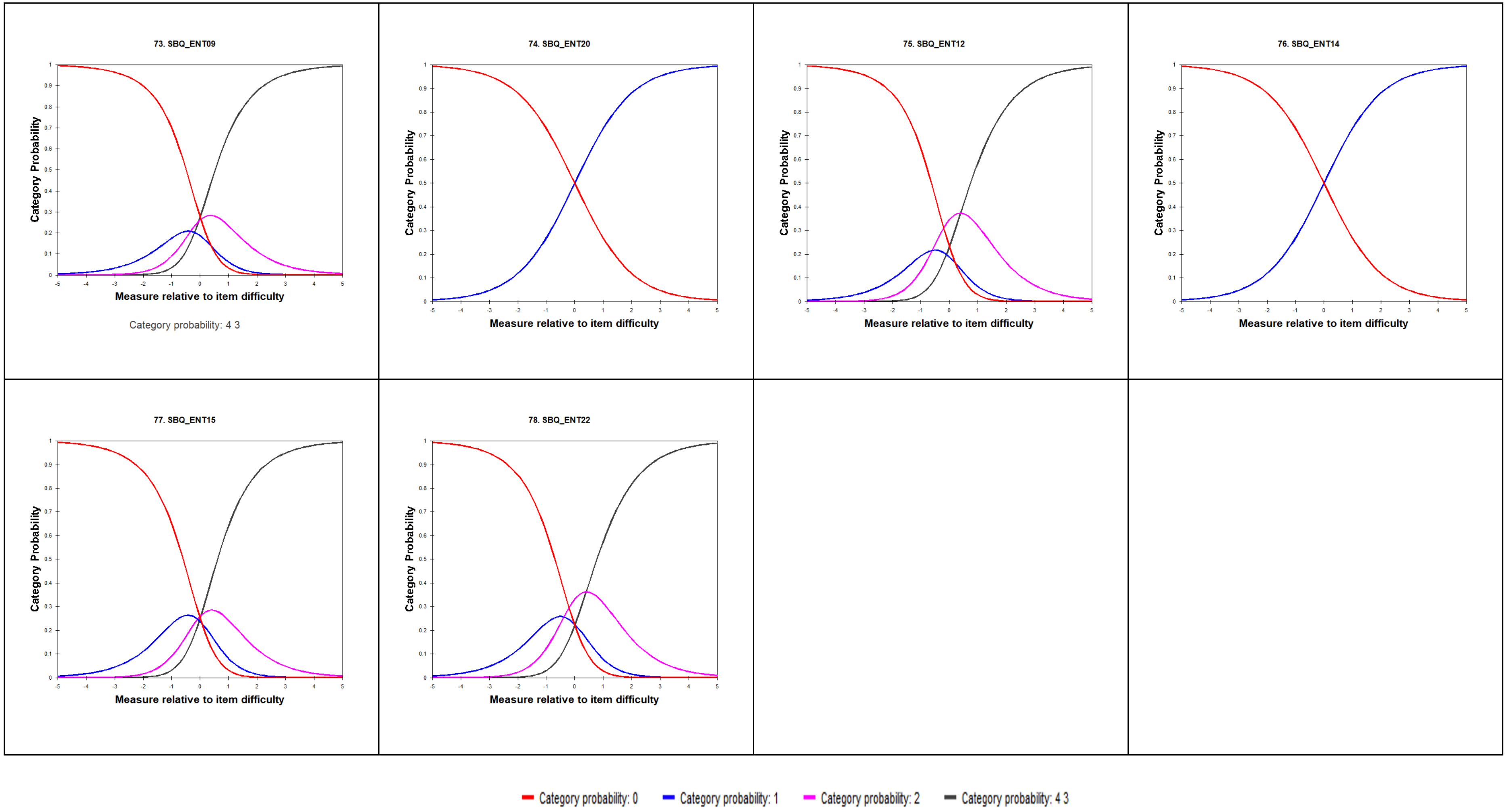

#### Category Probability Curves – Stomach and Digestion

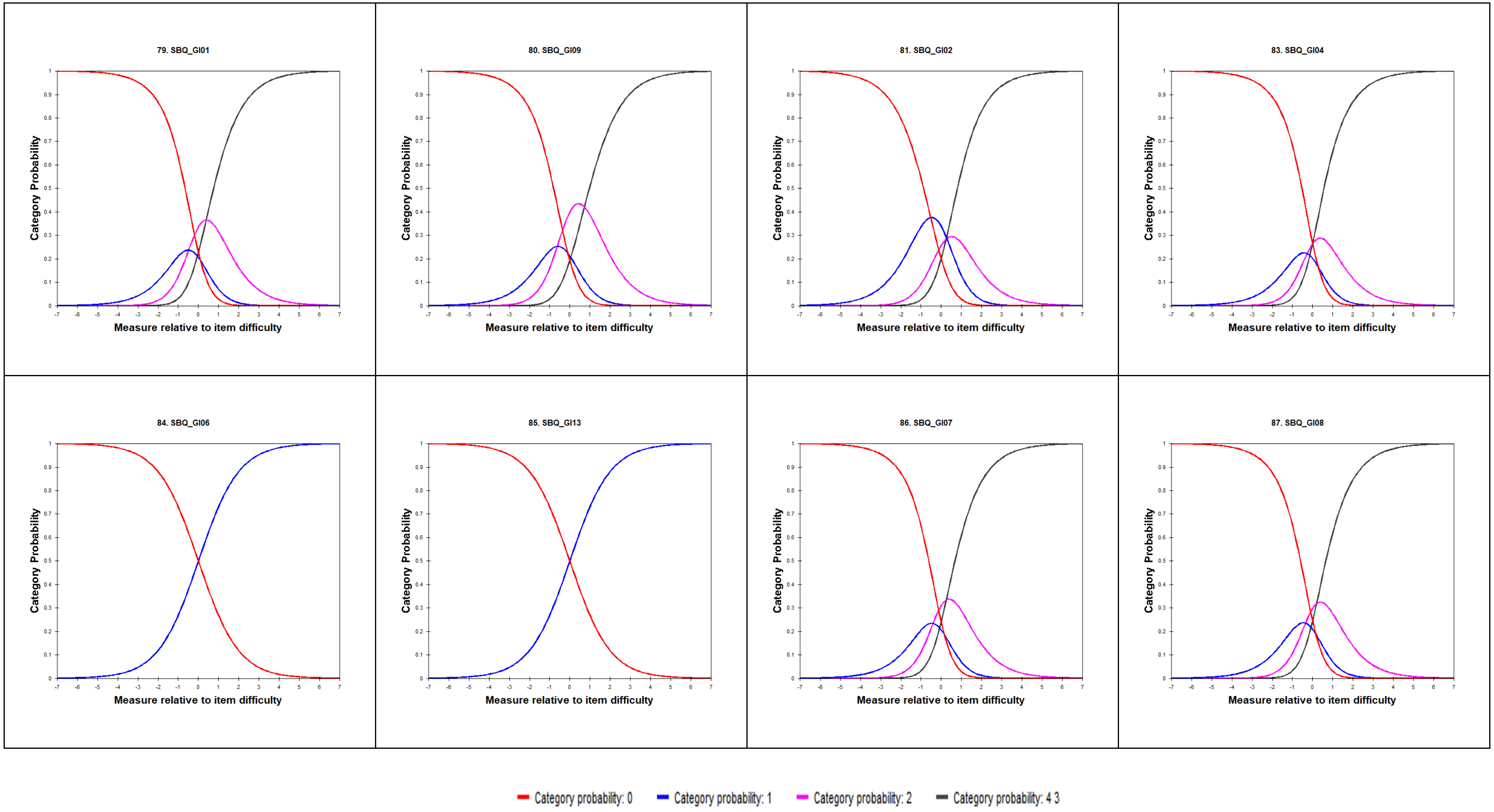

### Category Probability Curves – Muscles and Joints

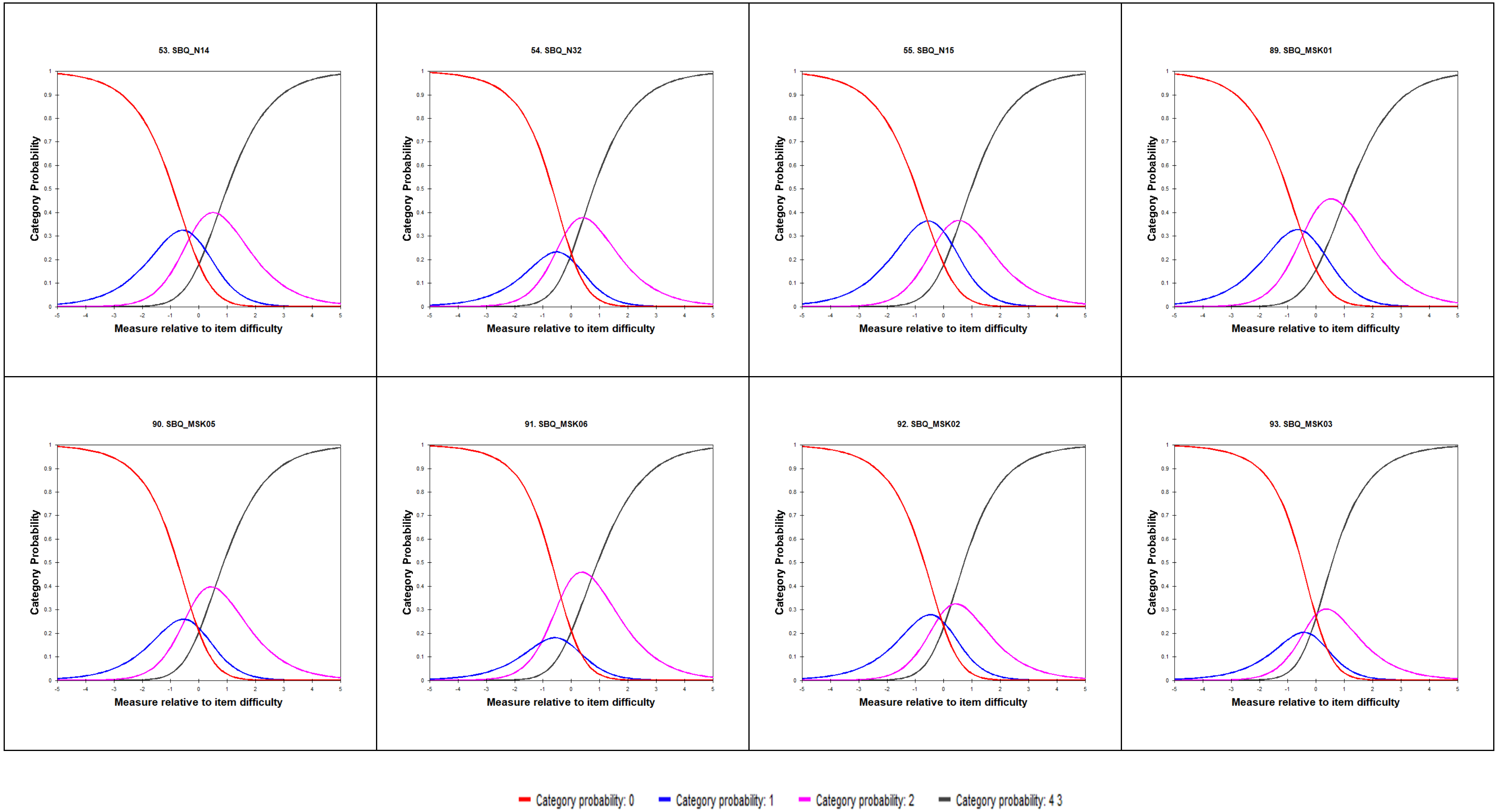

#### Category Probability Curves – Muscles and Joints continued...

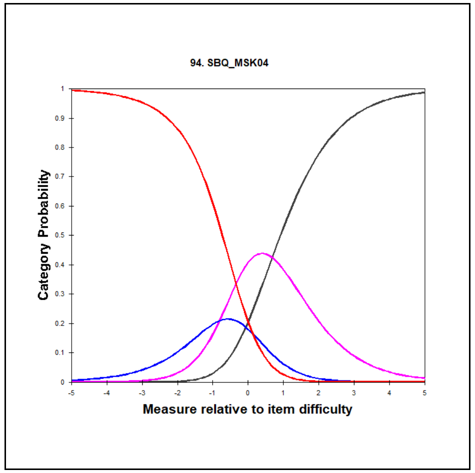


Category probability: 0 Category probability: 1 Category probability: 2 Category probability: 4 3

### Category Probability Curves – Mental Health and Wellbeing

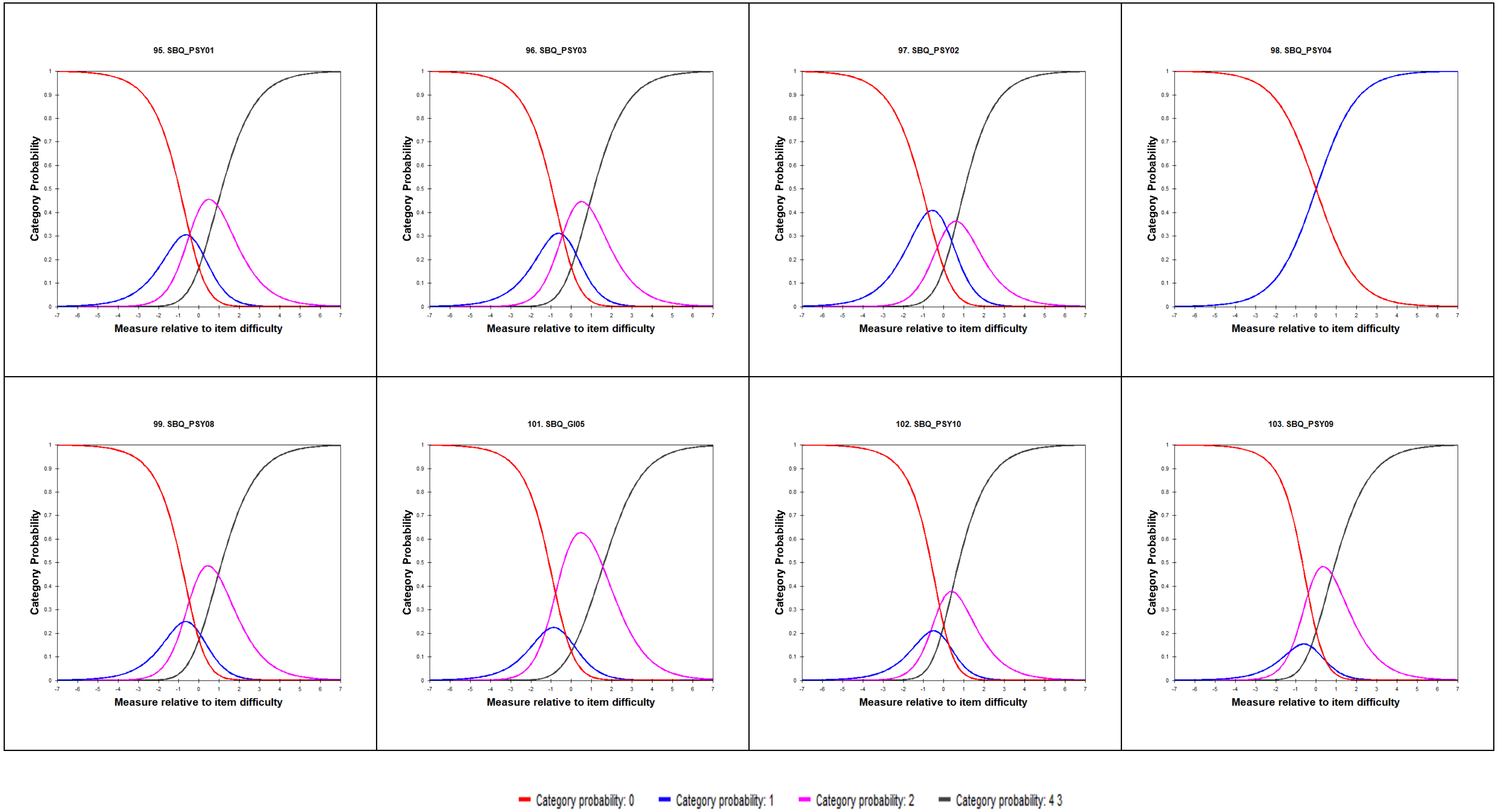

#### Scale: Mental Health and Wellbeing continued...

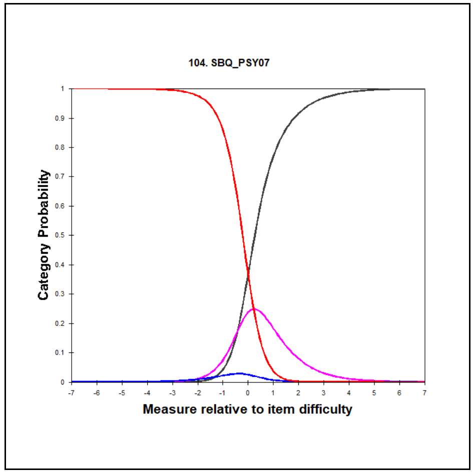


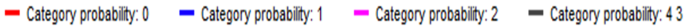

### Category Probability Curves – Skin and Hair

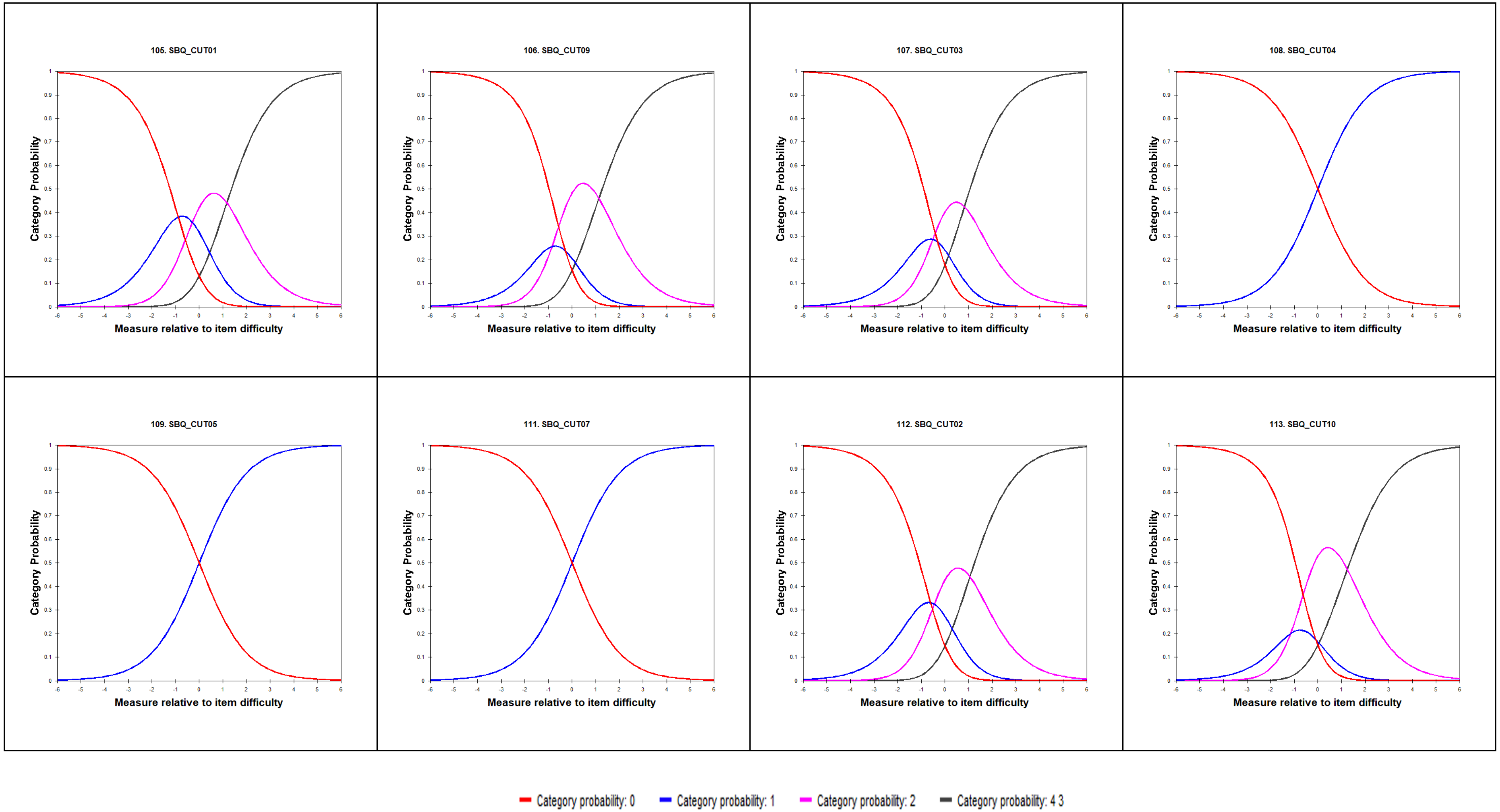

### Category Probability Curves – Eyes

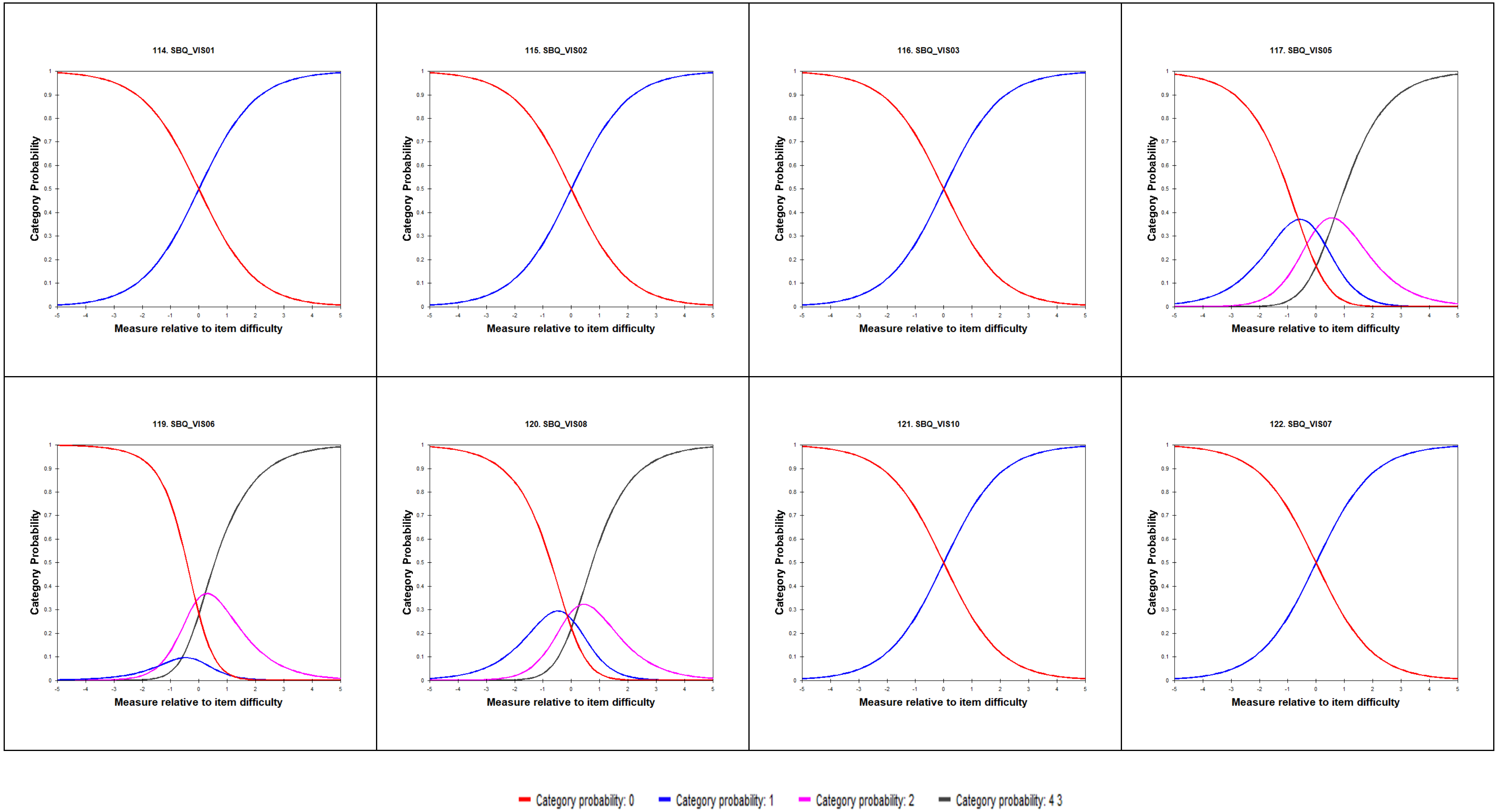

### Category Probability Curves – Eyes

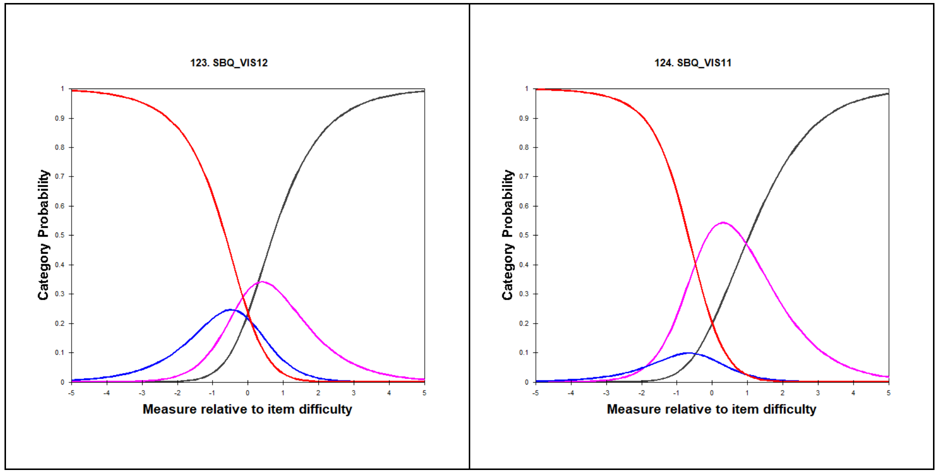


Category probability: 0 Category probability: 1 Category probability: 2 Category probability: 4 3

### Category Probability Curves – Female Reproductive and Sexual Health

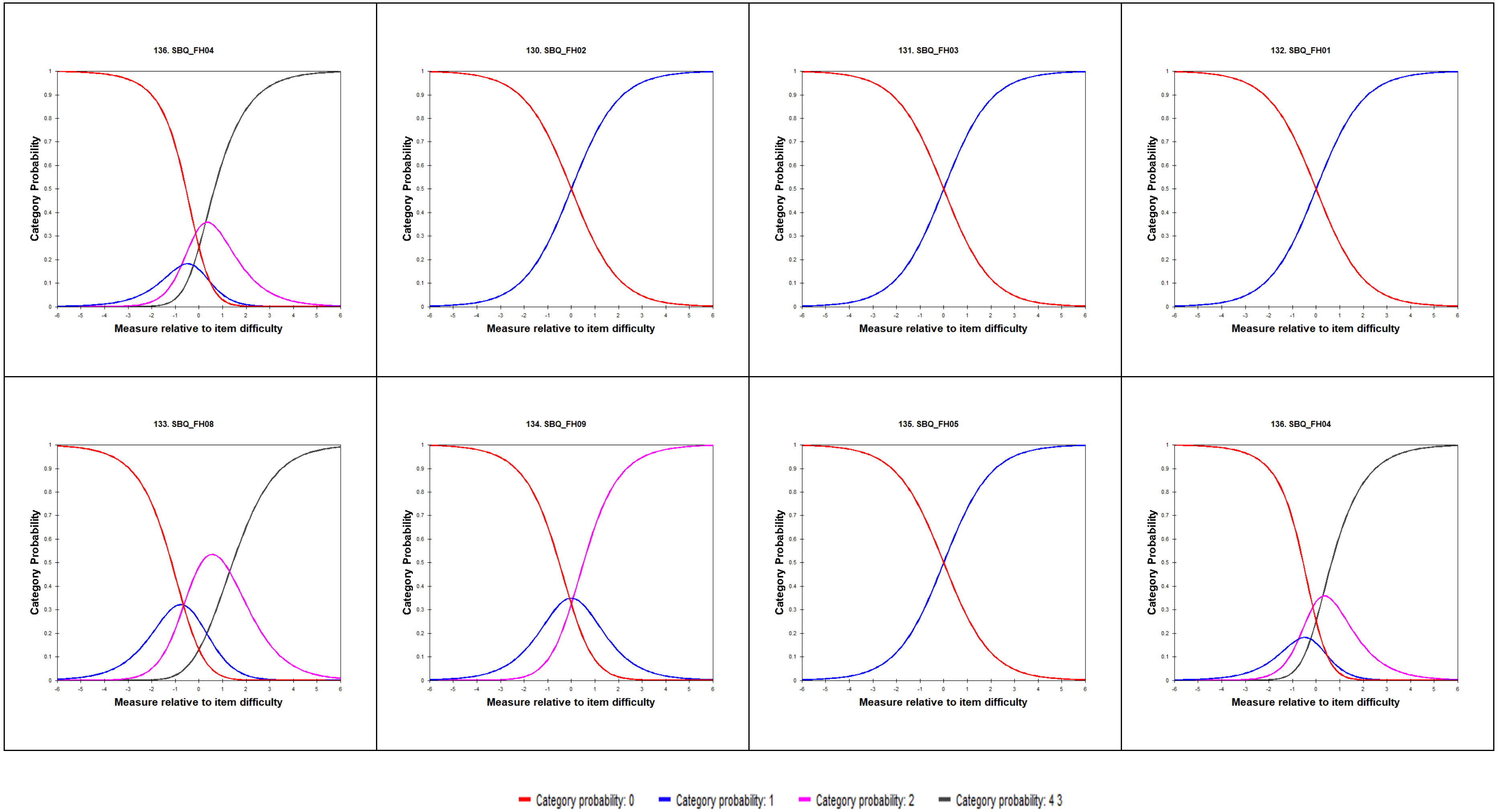

### Category Probability Curves – Male Reproductive and Sexual Health

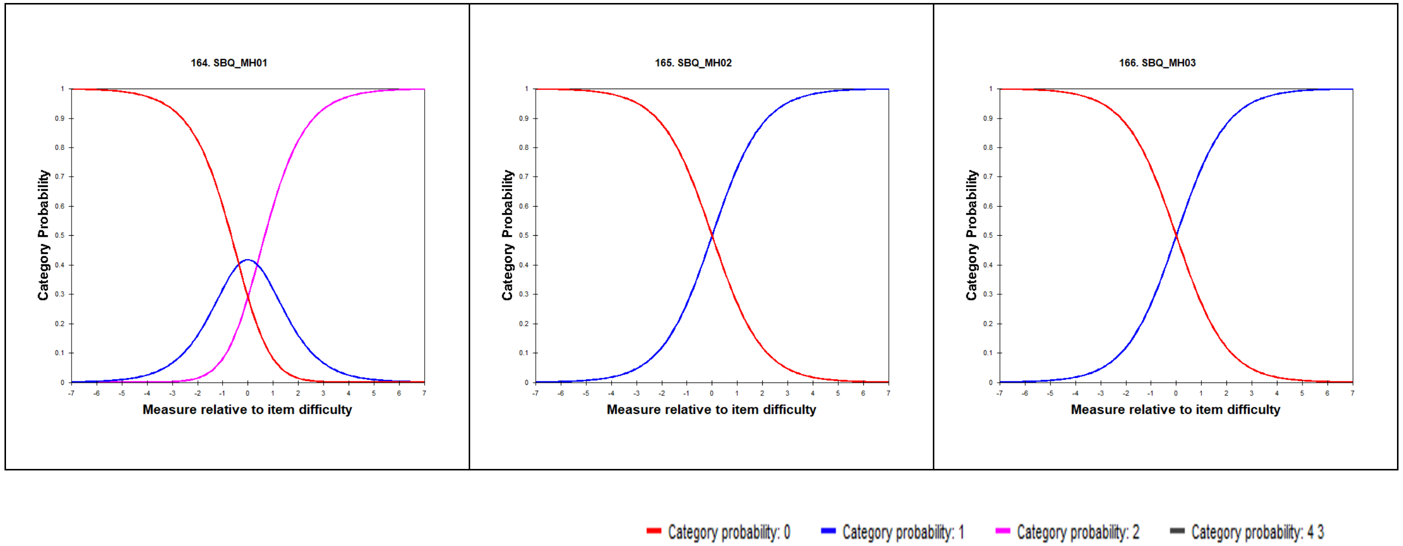

### Category Probability Curves – Other Symptoms

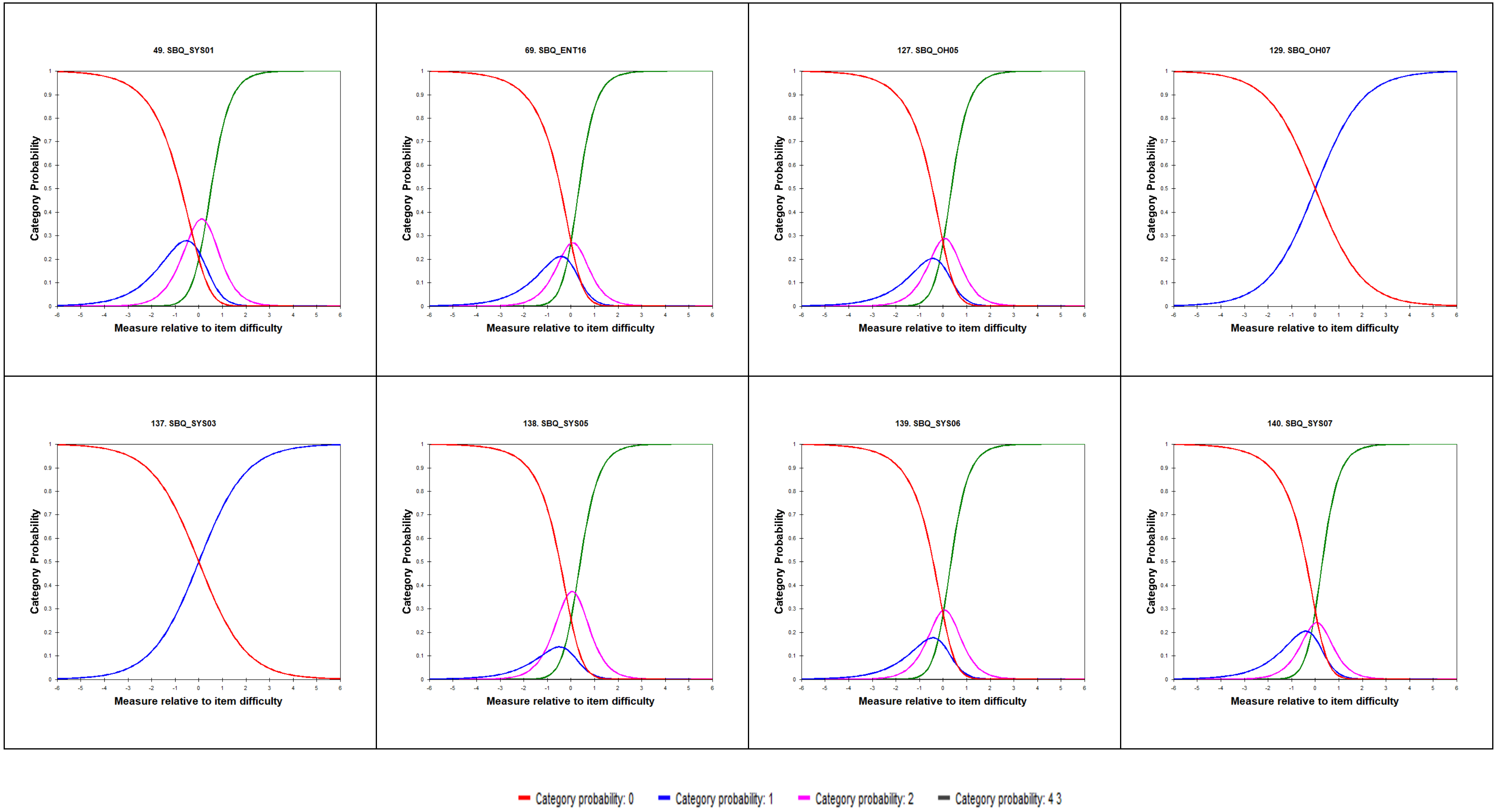

#### Category Probability Curves – Other symptoms continued...

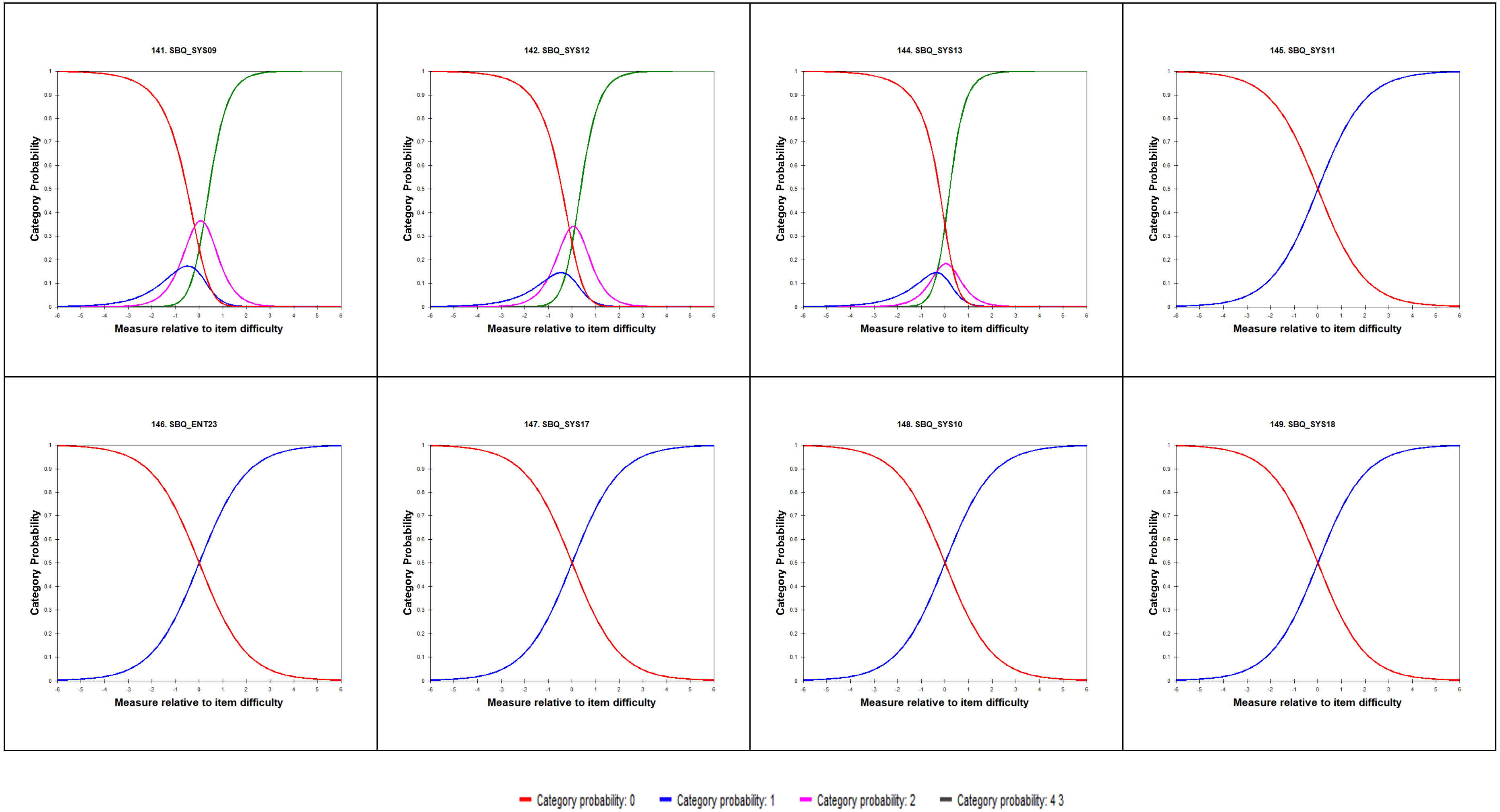

### Category Probability Curves – Other symptoms continued...

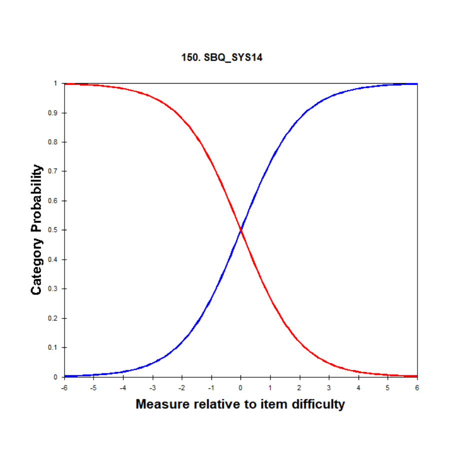


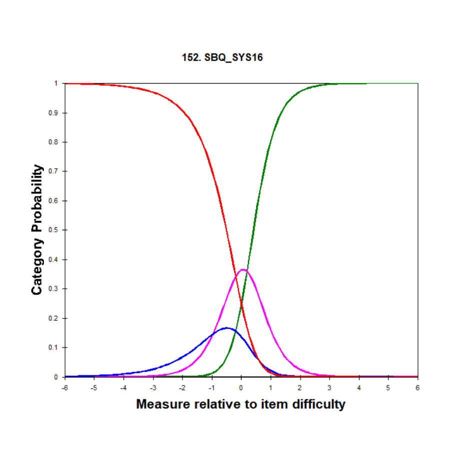


Category probability: 0 Category probability: 1 Category probability: 2 Category probability: 4 3

### Category Probability Curves – Interference

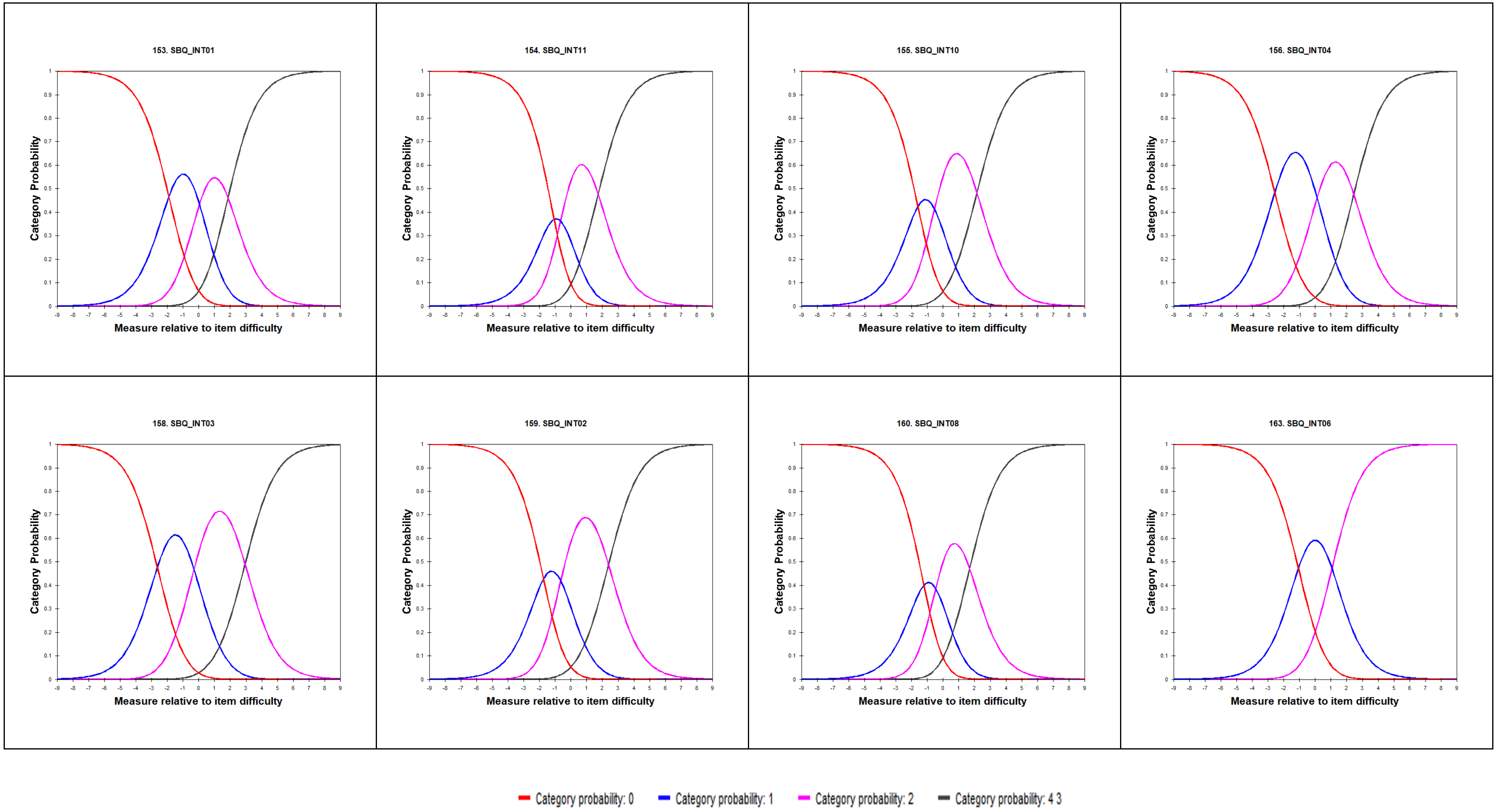
