## Supplementary material for "Development and validation of the Symptom Burden Questionnaire™ for Long Covid: A Rasch analysis": S2 Figure

### S2 Figure: Item-Person Maps for the SBQ™-LC (Version 1.0)

For each scale, the person-item map displays the location of person abilities and item difficulties respectively along the same latent dimension (y-axis).

**Scale: Breathing**

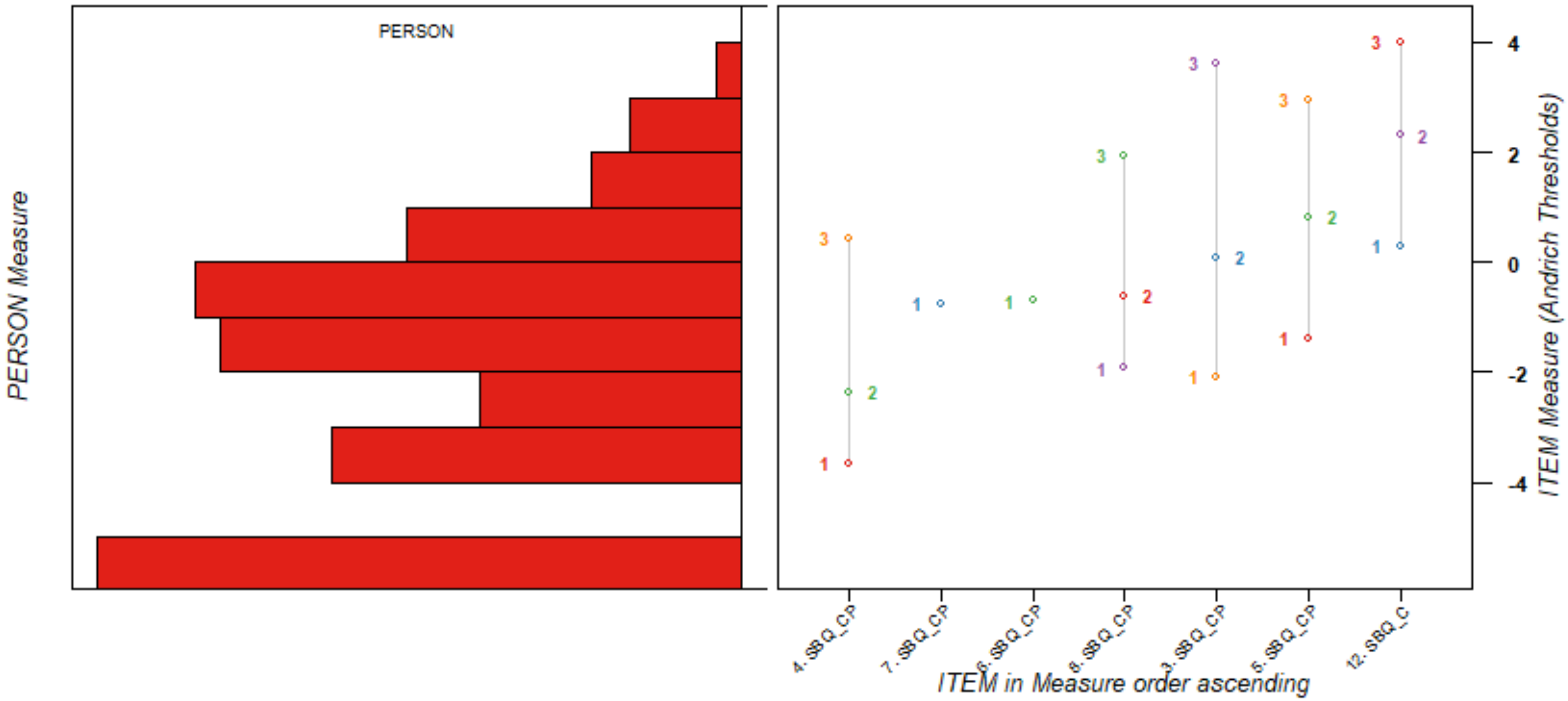

**Scale: Pain**

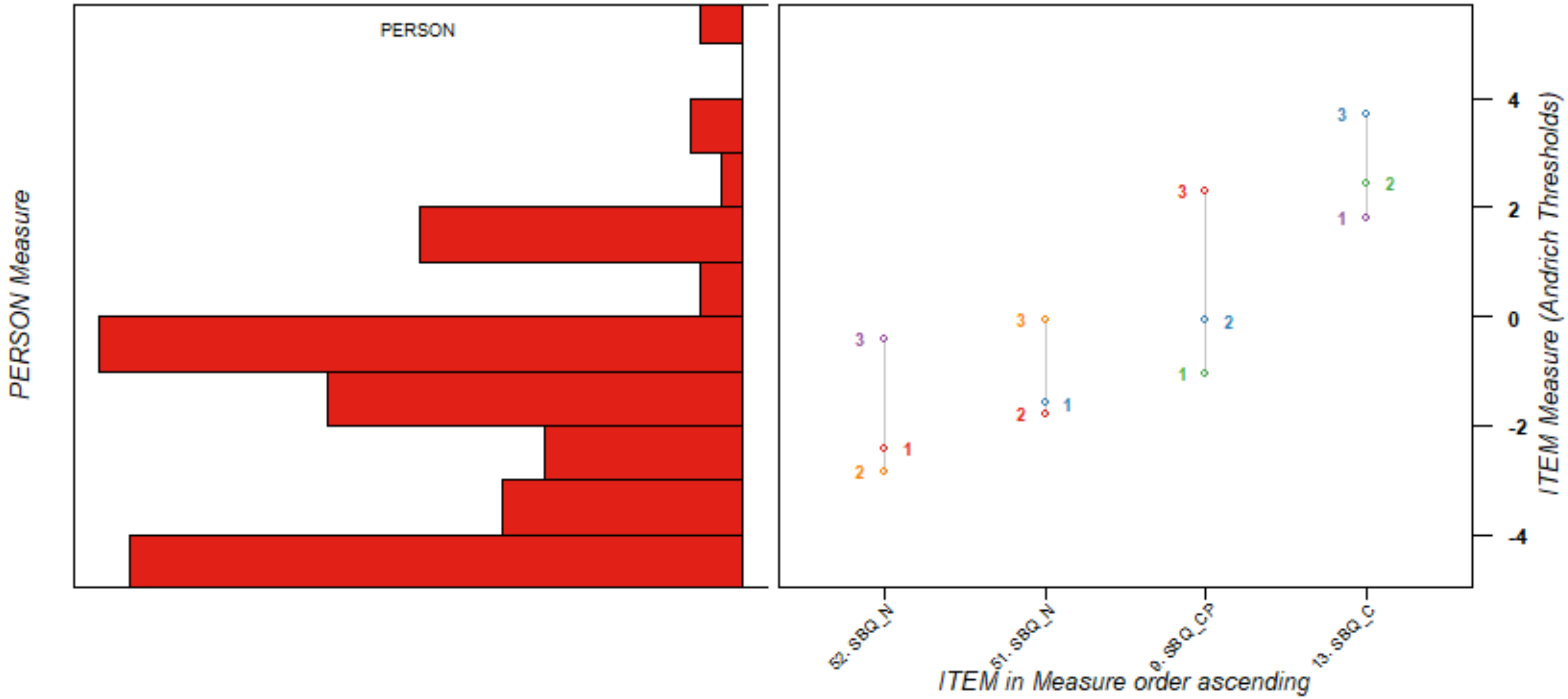
PERSON
PERSON Measure
ITEM Measure (Andrich Thresholds)
4
2
0
-2
-4
52. SBQ_N
51. SBQ_N
9. SBQ_CP
13. SBQ_C
ITEM in Measure order ascending

**Scale: Circulation**

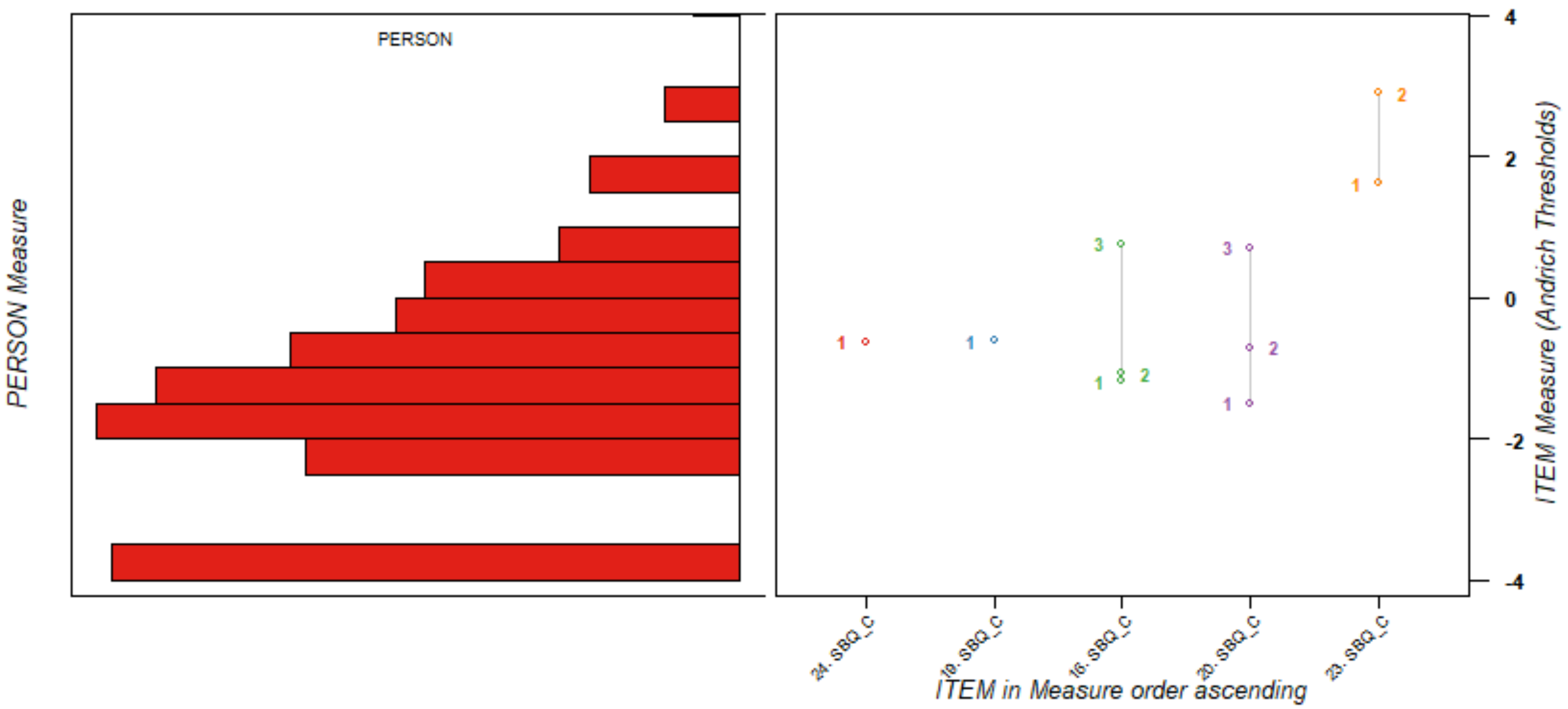

Scale: Fatigue

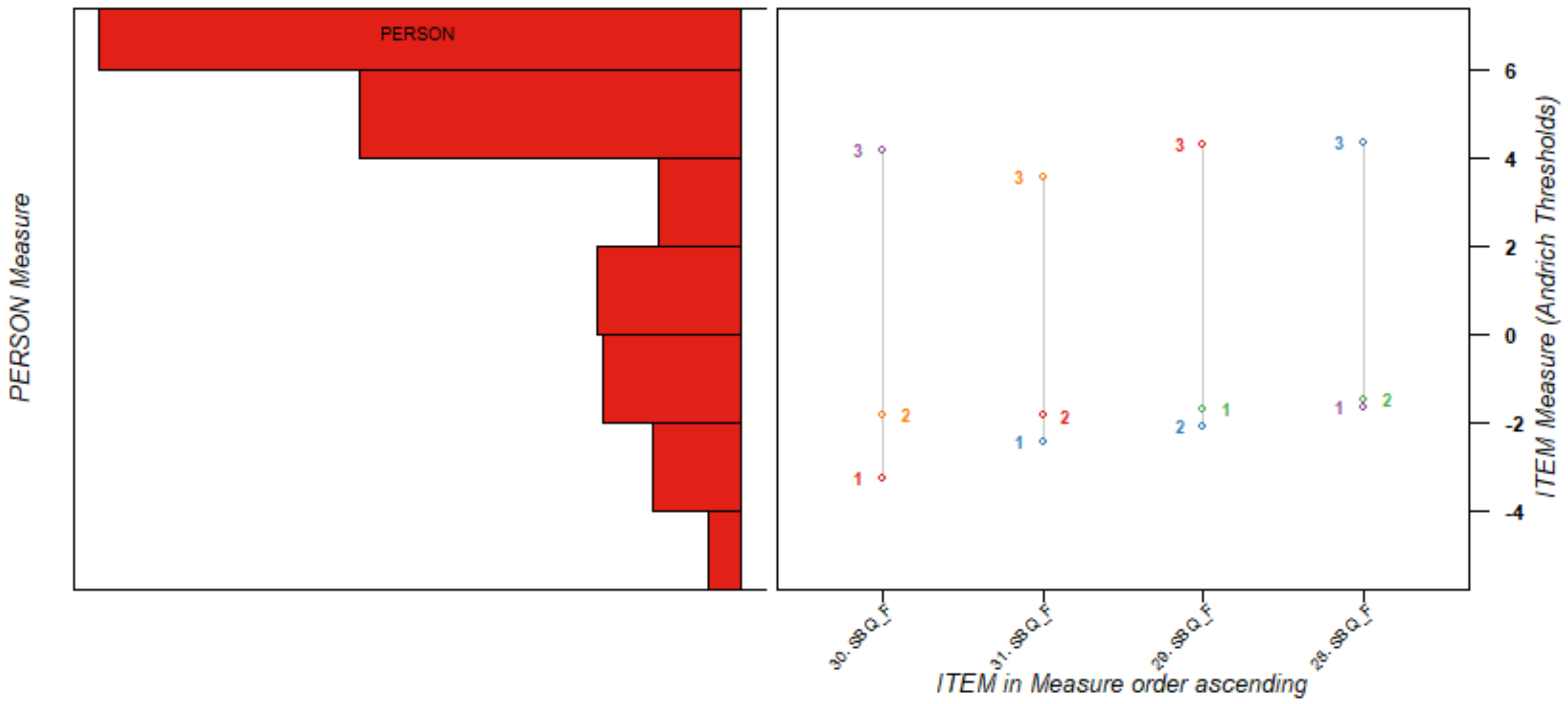

**Scale: Memory, Thinking & Communication**

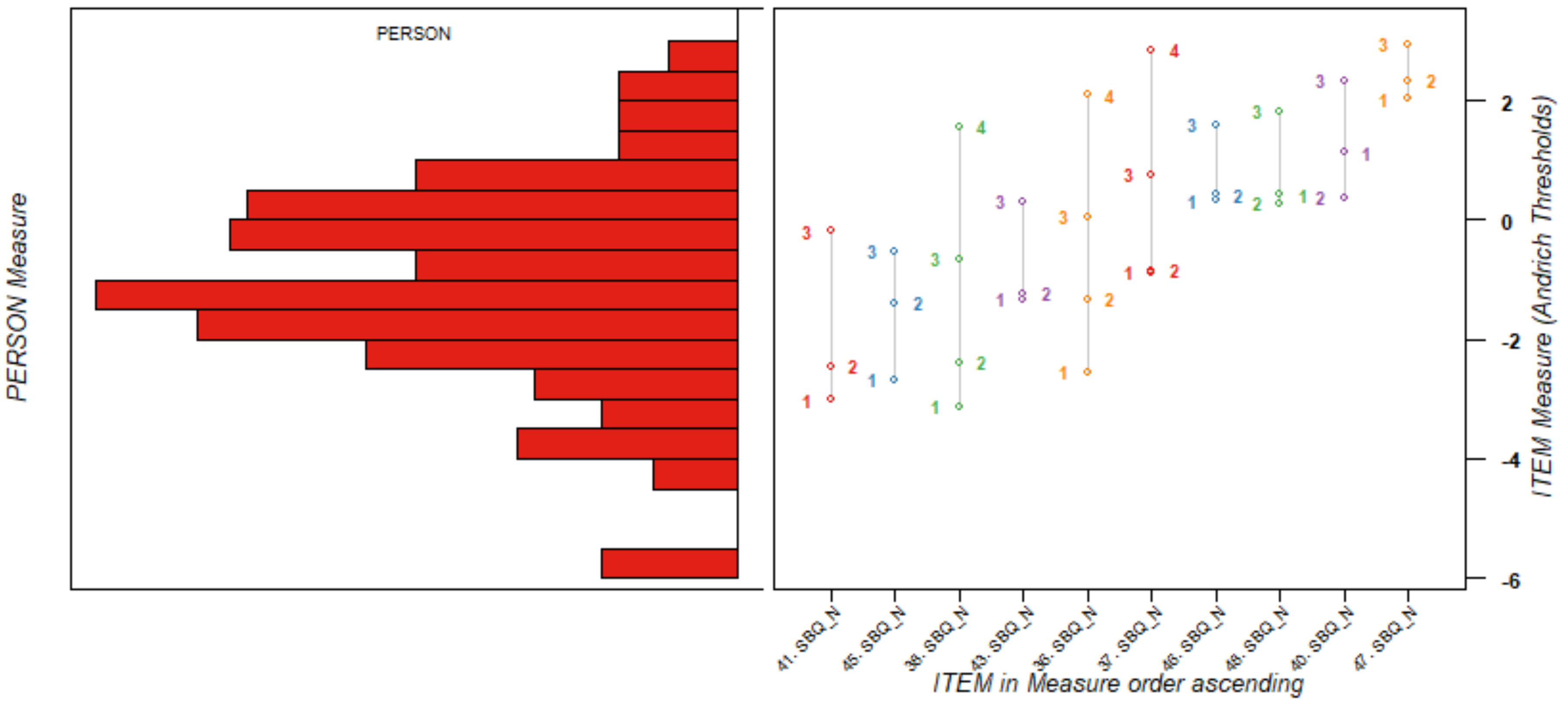

**Scale: Movement**

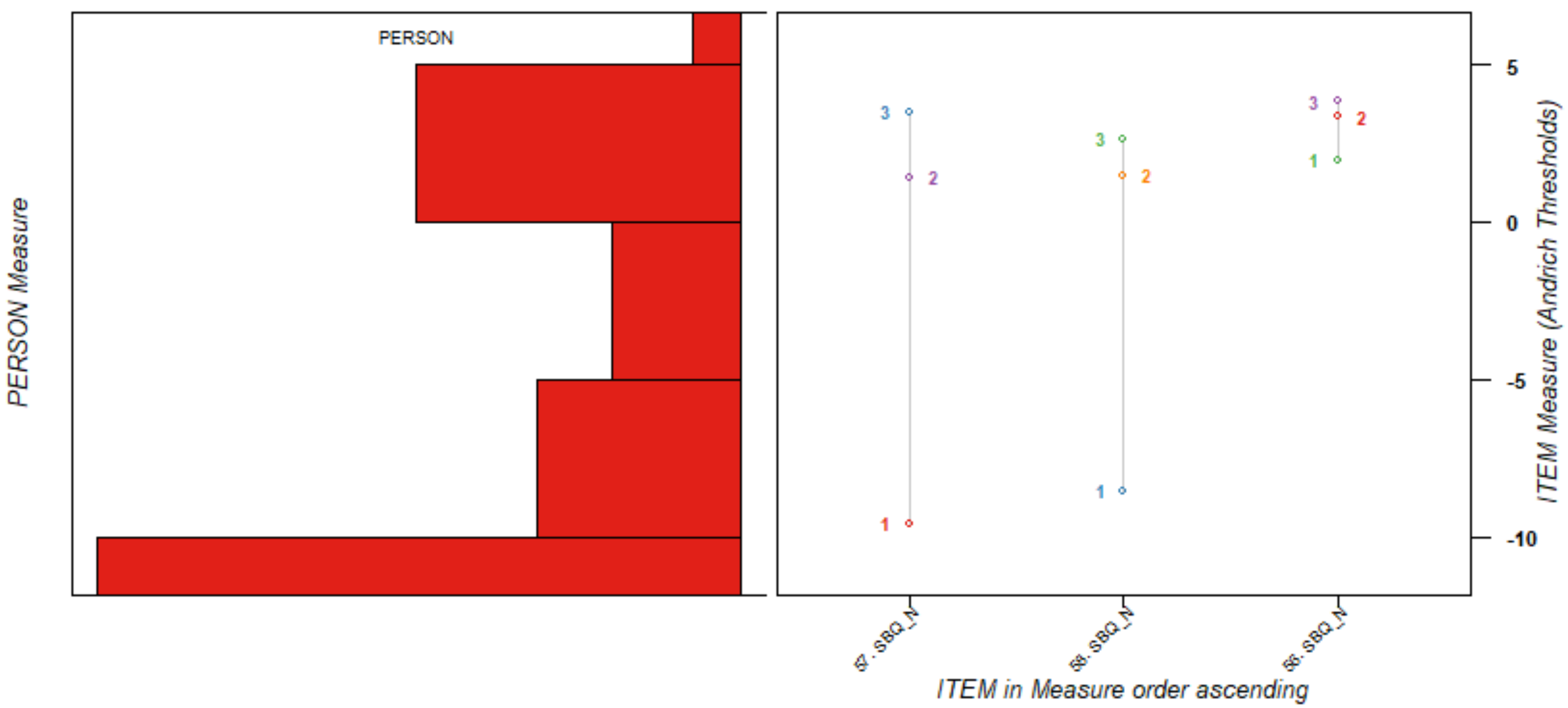

**Scale: Sleep**

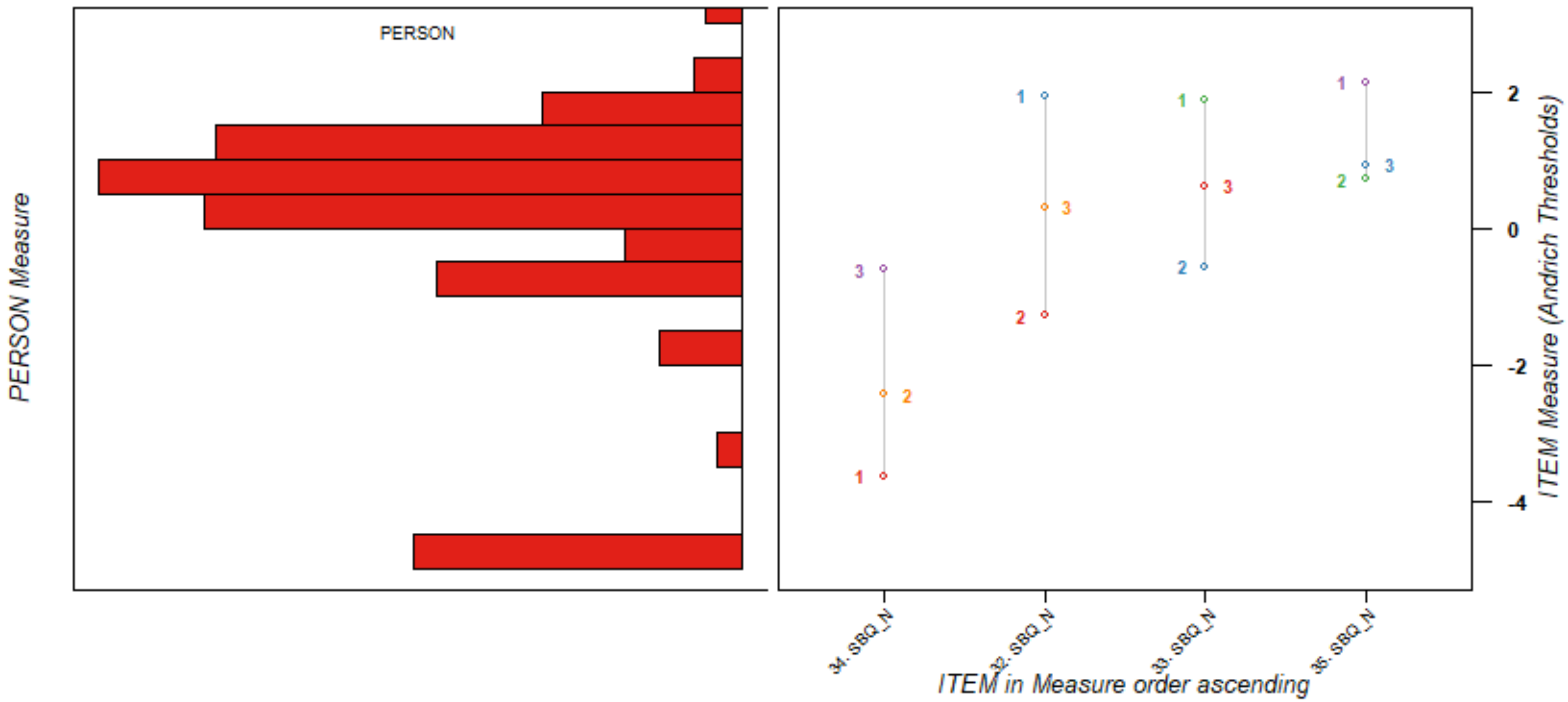
PERSON
PERSON Measure
ITEM Measure (Andrich Thresholds)
2
0
-2
-4
34. SBQ_N
32. SBQ_N
33. SBQ_N
35. SBQ_N
ITEM in Measure order ascending

**Scale: Ear, Nose & Throat**

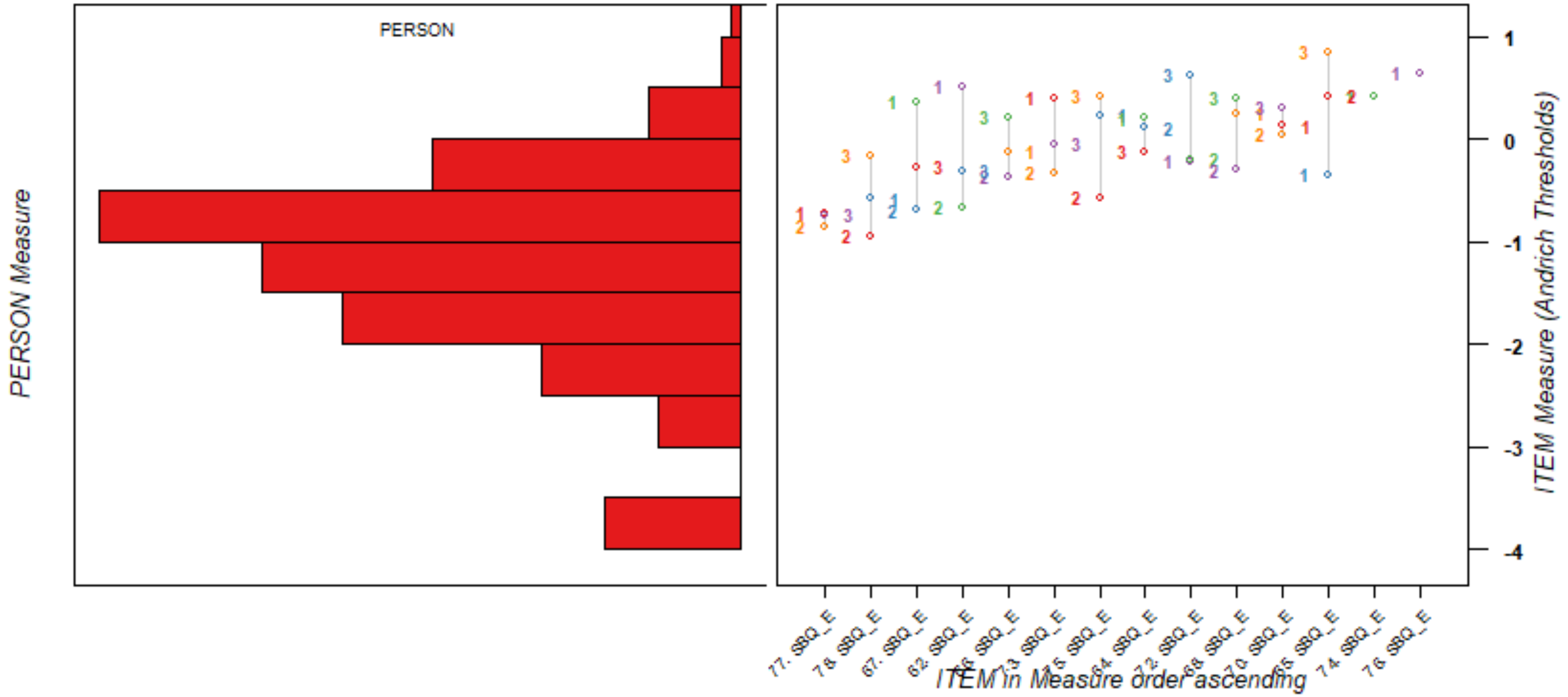

**Scale: Stomach and Digestion**

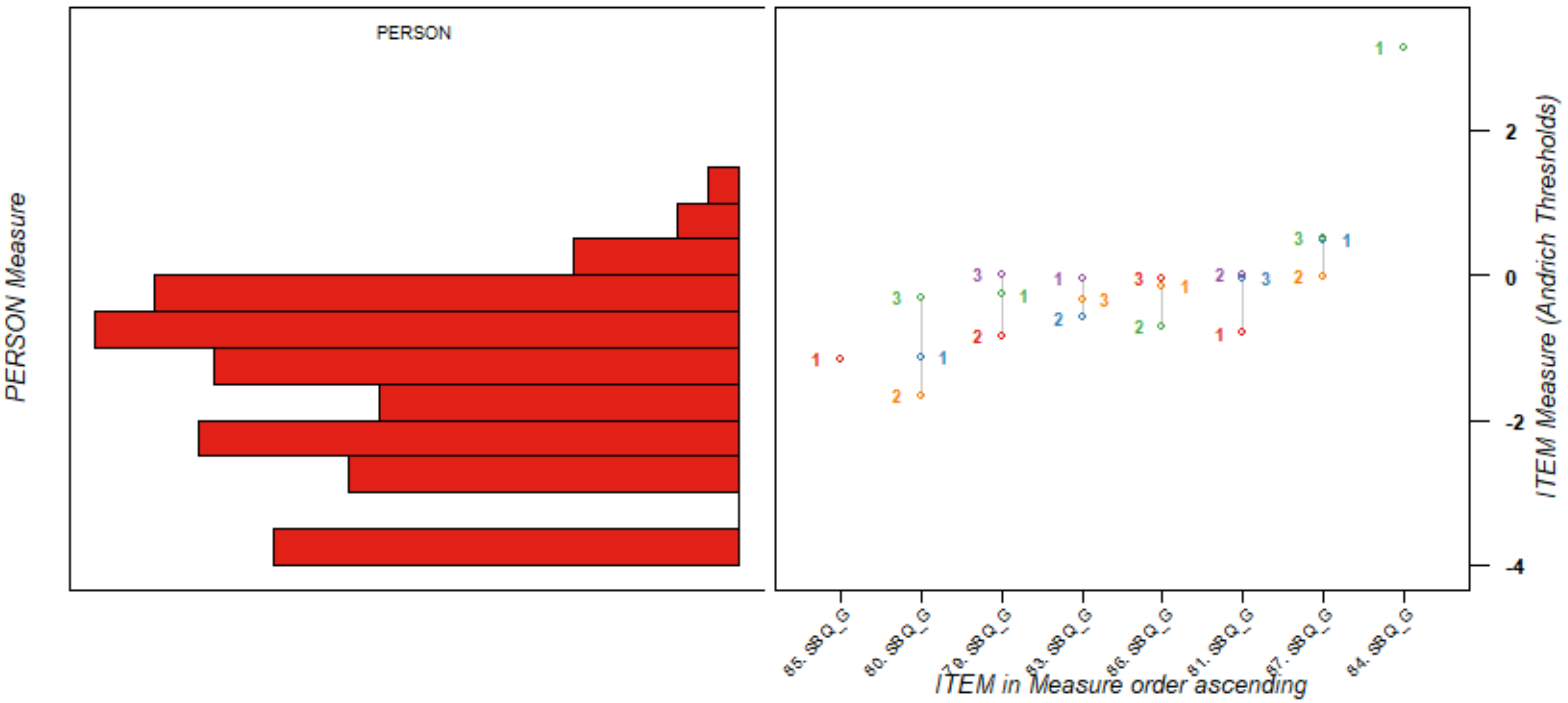

**Item-Person Map**
**Scale: Muscles and Joints**

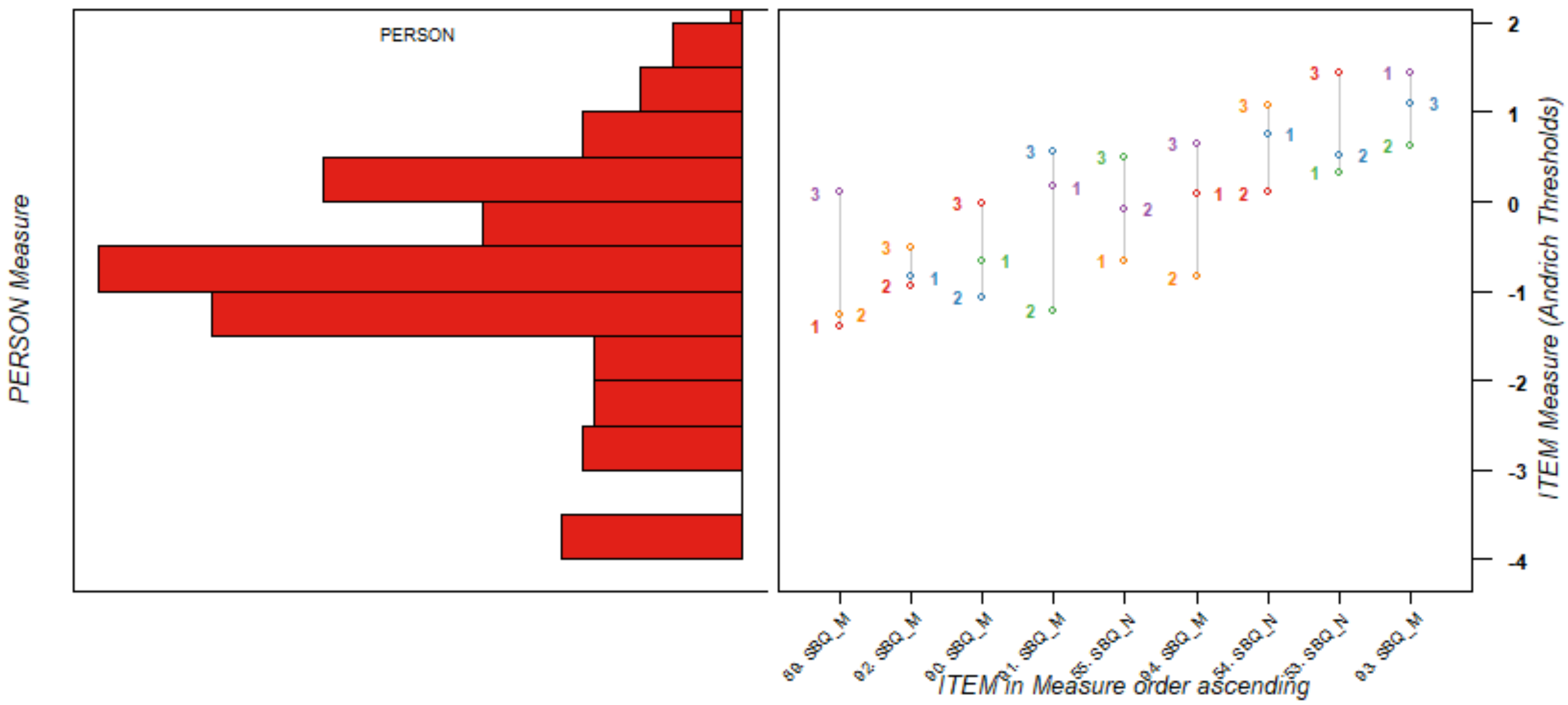

**Scale: Mental Health & Wellbeing**

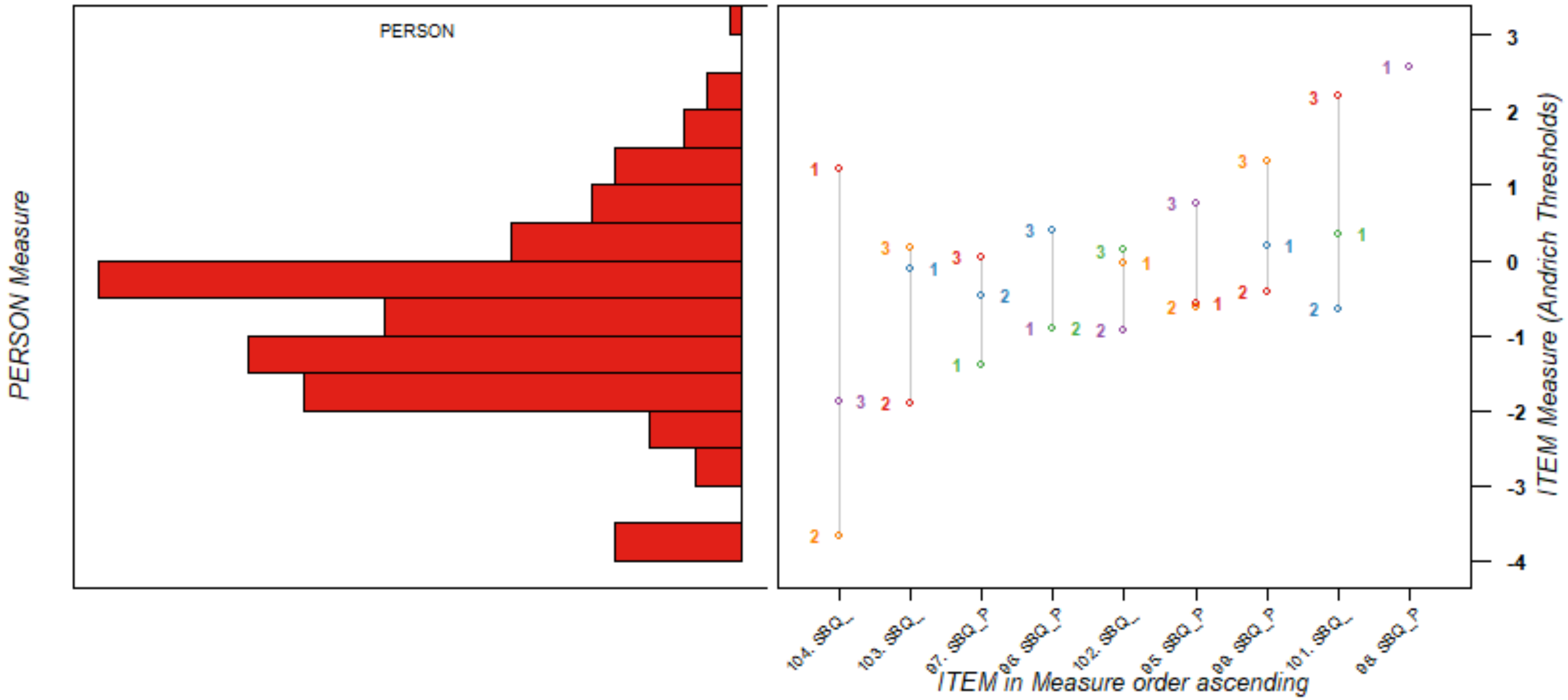

**Scale: Skin and Hair**

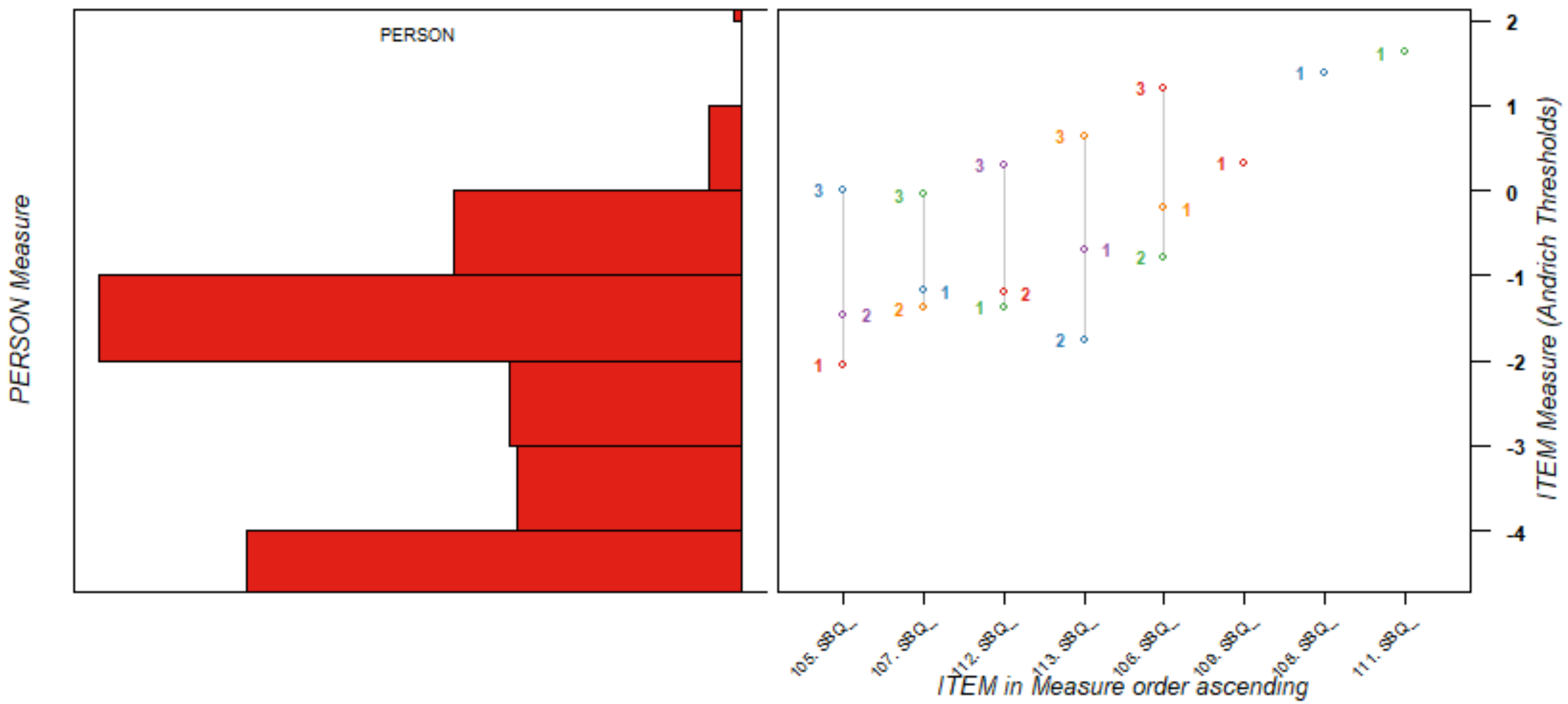

**Scale: Eyes**

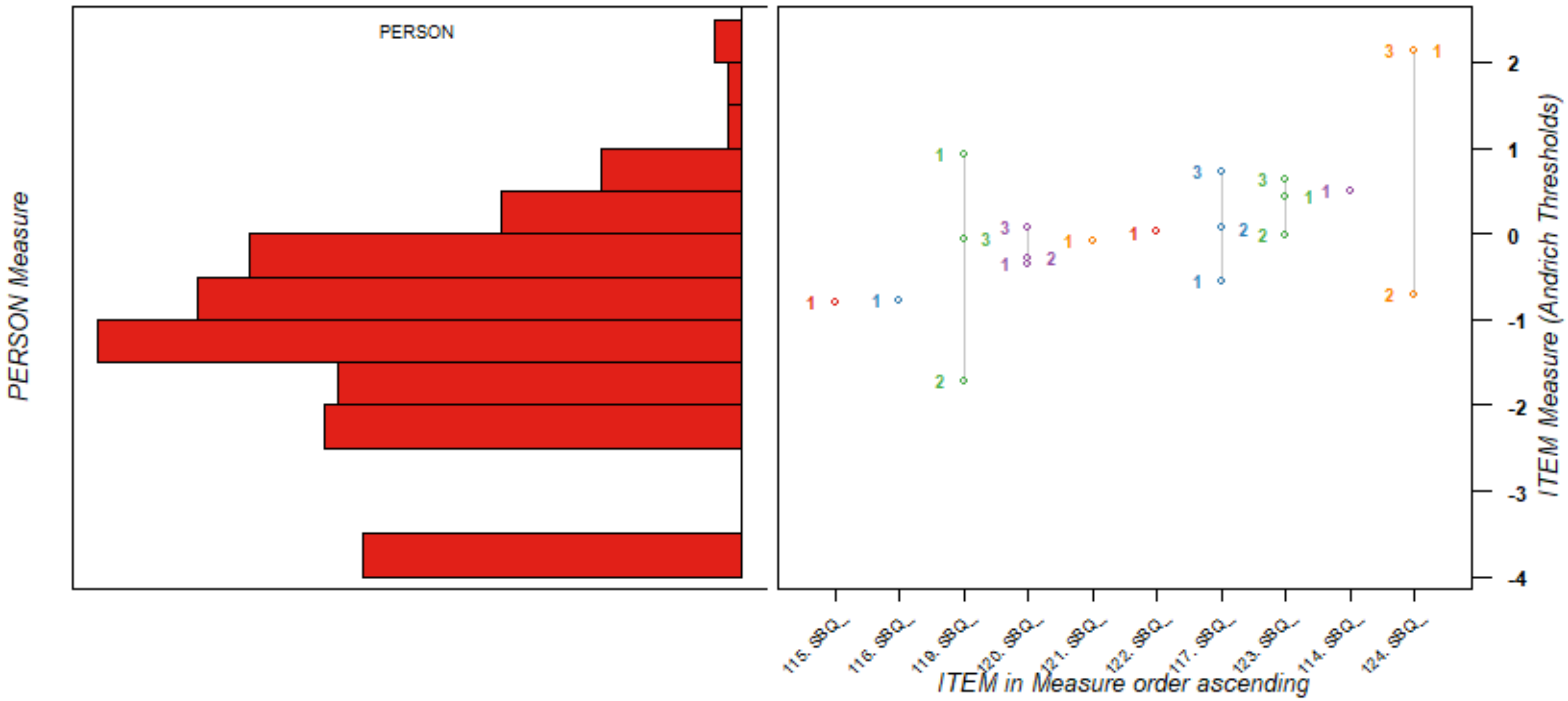
PERSON
PERSON Measure
ITEM Measure (Andrich Thresholds)
ITEM in Measure order ascending
115. SBQ_
116. SBQ_
119. SBQ_
120. SBQ_
121. SBQ_
122. SBQ_
117. SBQ_
123. SBQ_
114. SBQ_
124. SBQ_
2
1
0
-1
-2
-3
-4

**Scale: Female Reproductive and Sexual Health**

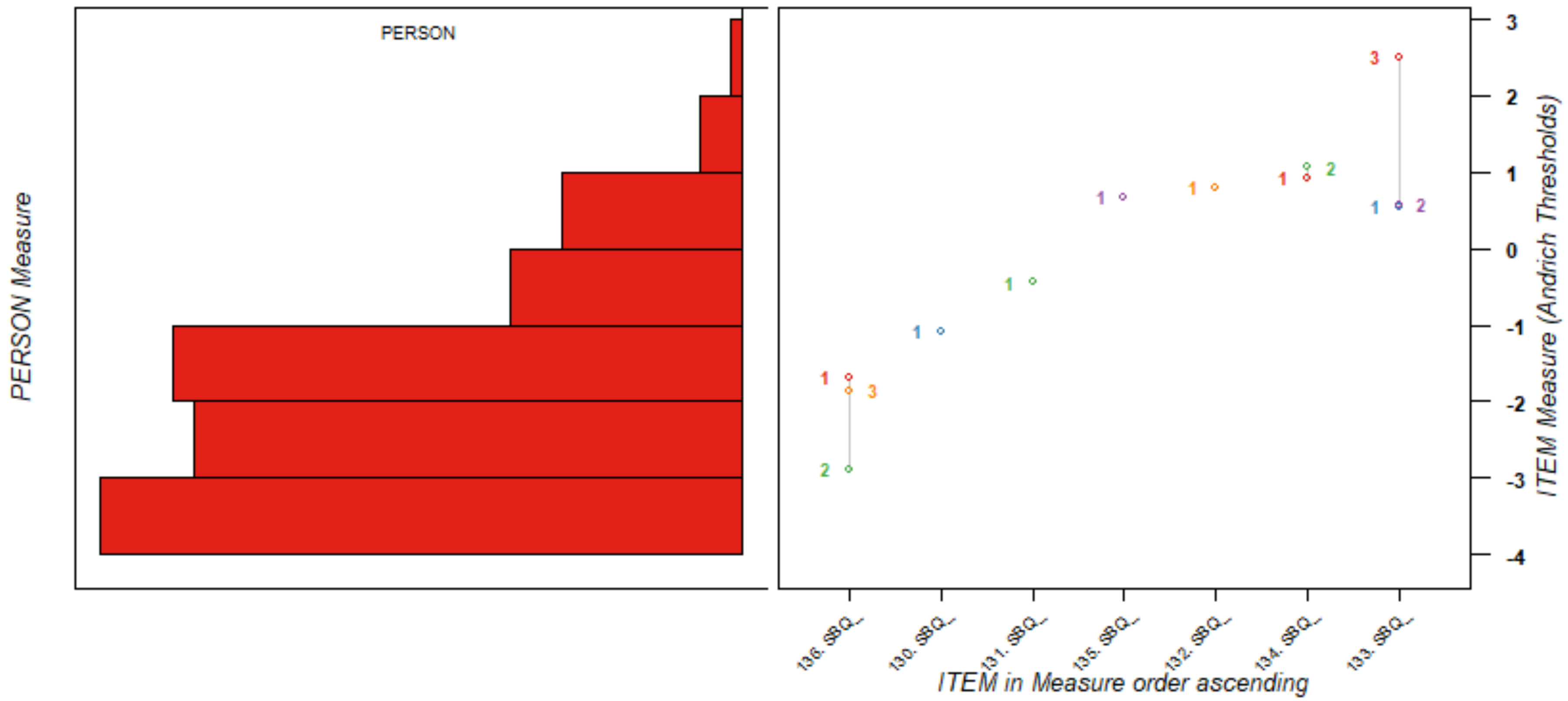

**Scale: Male Reproductive and Sexual Health**

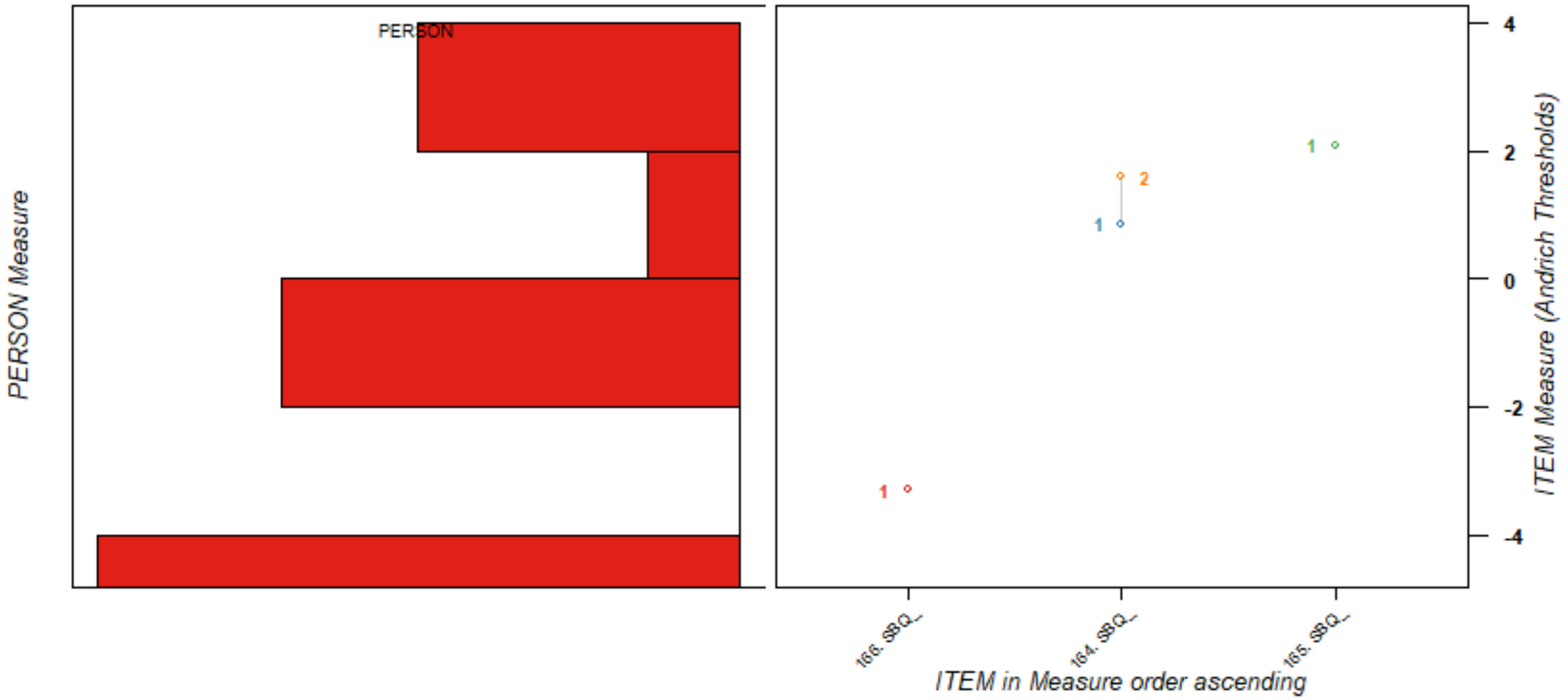

**Scale: Other Symptoms**

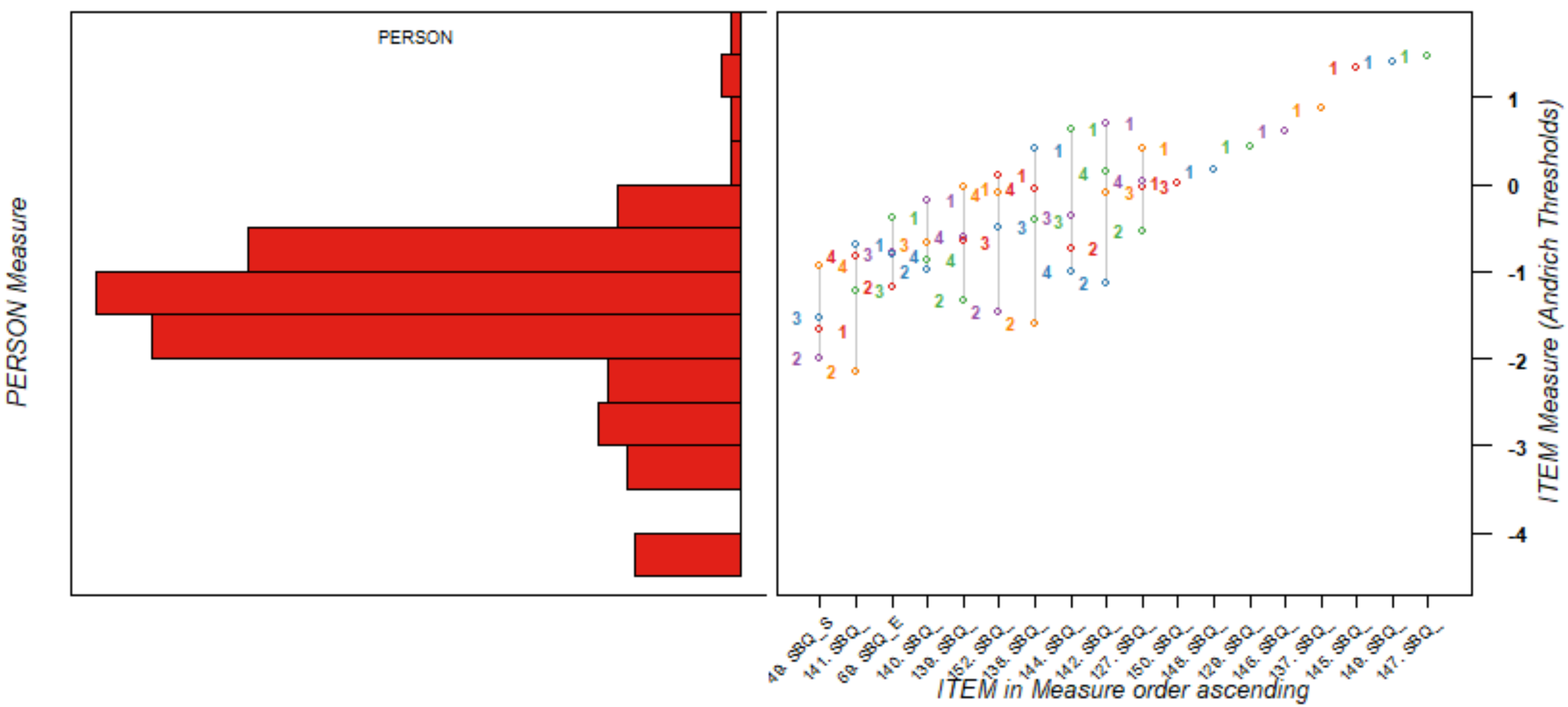

**Scale: Interference**

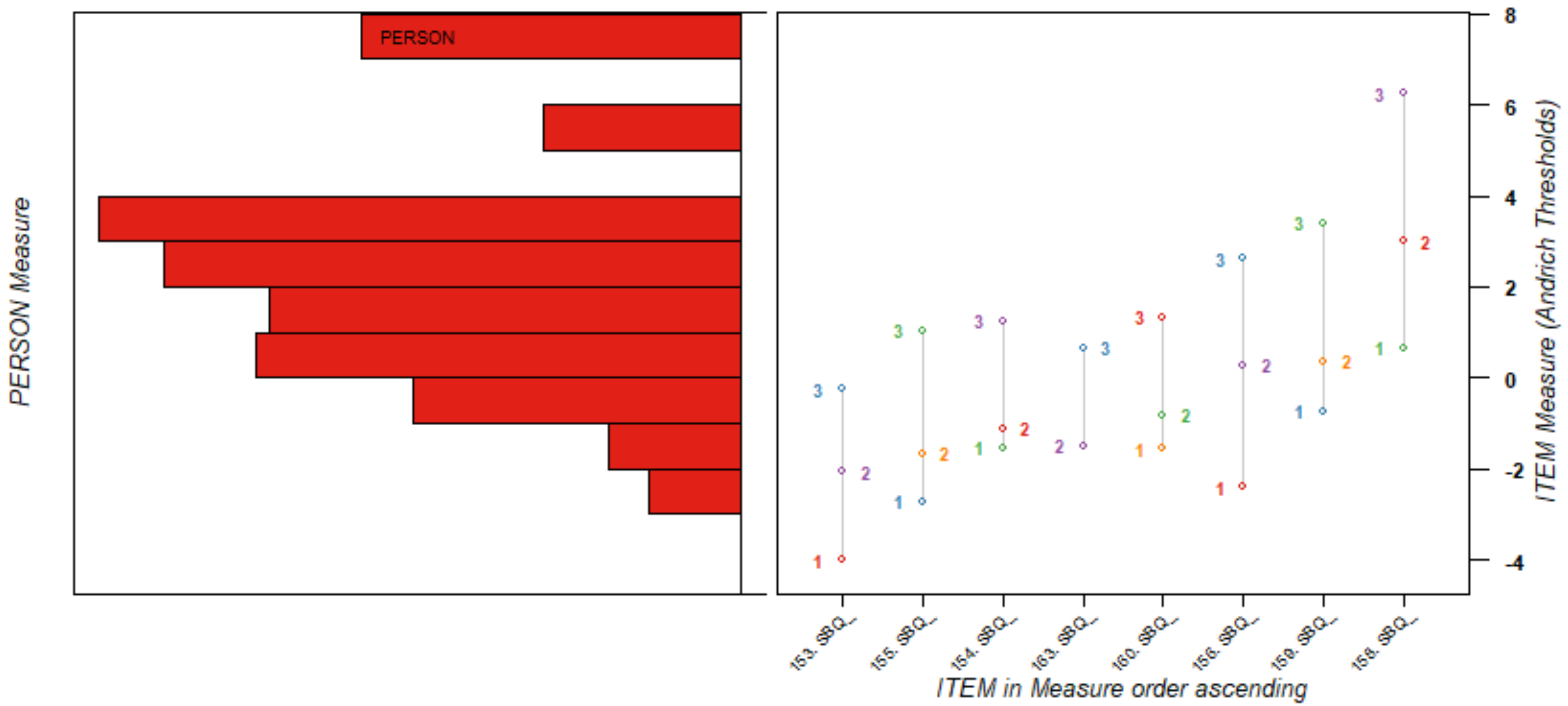
